# A scalable variational approach to characterize pleiotropic components across thousands of human diseases and complex traits using GWAS summary statistics

**DOI:** 10.1101/2023.03.27.23287801

**Authors:** Zixuan Zhang, Junghyun Jung, Artem Kim, Noah Suboc, Steven Gazal, Nicholas Mancuso

## Abstract

Genome-wide association studies (GWAS) across thousands of traits have revealed the pervasive pleiotropy of trait-associated genetic variants. While methods have been proposed to characterize pleiotropic components across groups of phenotypes, scaling these approaches to ultra large-scale biobanks has been challenging. Here, we propose FactorGo, a scalable variational factor analysis model to identify and characterize pleiotropic components using biobank GWAS summary data. In extensive simulations, we observe that FactorGo outperforms the state-of-the-art (model-free) approach tSVD in capturing latent pleiotropic factors across phenotypes, while maintaining a similar computational cost. We apply FactorGo to estimate 100 latent pleiotropic factors from GWAS summary data of 2,483 phenotypes measured in European-ancestry Pan-UK BioBank individuals (N=420,531). Next, we find that factors from FactorGo are more enriched with relevant tissue-specific annotations than those identified by tSVD (P=2.58E-10), and validate our approach by recapitulating brain-specific enrichment for BMI and the height-related connection between reproductive system and muscular-skeletal growth. Finally, our analyses suggest novel shared etiologies between rheumatoid arthritis and periodontal condition, in addition to alkaline phosphatase as a candidate prognostic biomarker for prostate cancer. Overall, FactorGo improves our biological understanding of shared etiologies across thousands of GWAS.

## Introduction

Genome-wide association studies (GWAS) have identified thousands of genetic variants that associate with complex traits and diseases affecting multiple traits^1–3^. Investigating this pervasive pleiotropy has enabled elucidating broader biological mechanisms, identifying comorbidity due to genetic susceptibility, and discovering or repurposing of therapeutic targets^4–6^.

Previous works have proposed methods to identify pleiotropic components under two related, but distinct camps of approaches. The first camp is to apply matrix factorization techniques (e.g., truncated singular value decomposition; tSVD) on a matrix of GWAS summary data^7–9^. While matrix factorization provides a computationally efficient means of capturing apparent pleiotropic components, its model-free approach leaves unclear what parameters are inferred from noisy observations (in this case effect-size estimates). The second camp of approaches are based on statistical models for genetic effects, but are limited to the analysis of a small number of traits due to computational demands^10–12^. As more GWAS summary data become available in large biobanks^13–15^, it is important to develop a scalable model-based approach that allows exploring the phenome-wide shared genetic architecture, either known or unknown to be genetically related *a priori*. Classical factor analysis provides an analogous approach towards summarizing shared latent factors in data, however inference in high-dimensional biobank settings is computationally demanding, thus limiting the scope of applied analysis.

Here, to identify latent pleiotropic components across thousands of phenotypes we present FactorGo, a **Factor** analysis model on **G**enetic ass**o**ciations using GWAS summary data. FactorGo models the uncertainty in genetic effect estimates and leverages an automatic relevance determination (ARD) prior to prune uninformative factors using a scalable variational Bayesian framework. Under extensive simulations, we find that FactorGo outperforms tSVD in reconstructing trait factor scores and is robust to model misspecifications. By analyzing thousands of phenotypes in Pan-UK Biobank, we identify alkaline phosphatase as a candidate prognostic biomarker for prostate cancer. Moreover, we recapitulate previously reported brain-specific enrichment for BMI and reproductive system and muscular-skeletal enrichment for height. For disease traits, we learn the shared bacterial etiology between rheumatoid arthritis and periodontal condition. Taken together, our results demonstrate that FactorGo prioritizes biologically meaningful latent pleiotropic factors which reflect shared biological mechanisms across traits.

## Material and Methods

### FactorGo model

Here, we briefly describe the FactorGo generative model of observed GWAS summary data assuming correlations in effects arise due to pleiotropy. For a full account please see details in **Supplemental Text 1**. In principle, FactorGo assumes the true genetic effect can be decomposed into latent pleiotropic factors (see **Figure 1**). Briefly, we model test statistics at *p* independent variants from the *i*^th^ GWAS 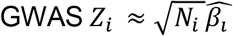 as a linear combination of *k* shared latent variant loadings *L* ∈ *R*^*p*×*k*^ with trait-specific factor scores *f*_*i*_ ∈ *R*^*k*×1^ as

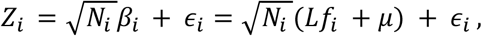

**Figure 1.**
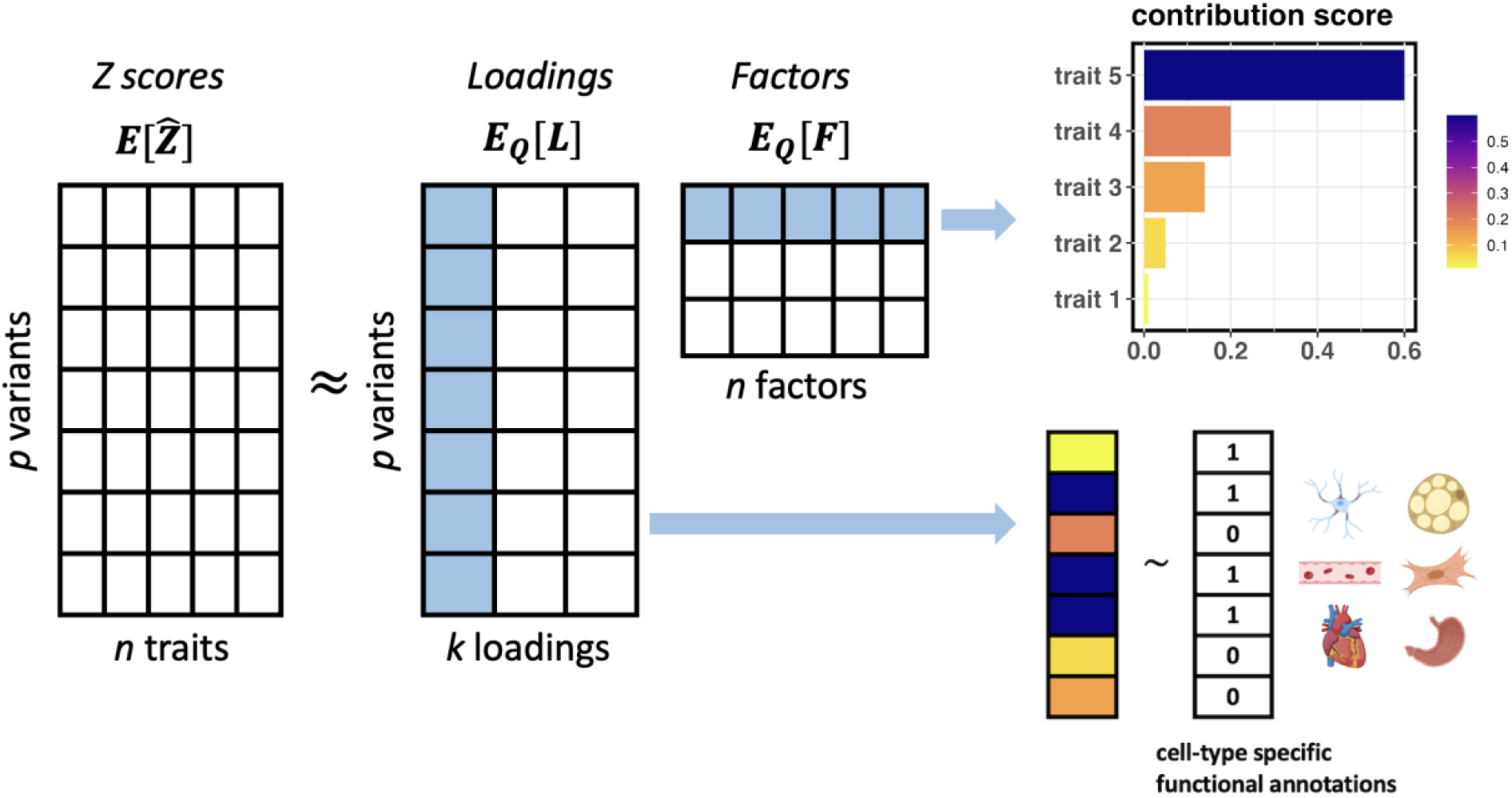
Overview of FactorGo. FactorGo decomposes the observed Z-score summary statistics of *p* variants in *n* traits to *k* pleiotropic factors. The column vector of *L* is variant loadings and row vector of *F* is the trait factor score for each inferred factor as highlighted in light blue. Here we plotted for *n* = 5, *p* = *7*, and *k* = *3* for illustrative purposes. To identify traits characterizing a given factor, we calculated contribution scores of this factore across all traits (top arrow). To understand the biological function of a given factor, we regressed transformed variant loadings on cell-type-specific annotations using LD score regression (bottom arrow). The colors on transformed scores represent the magnitude of values.

where *N*_*i*_ is the sample size for the *i*^th^ GWAS, *µ* is the intercept and *∈*_*i*_ *∼ N*(0, *τ*^*−*1^*I*_*p*_) reflects residual heterogeneity in statistical power across studies with precision scalar *τ*. Given *Z* = {*Z*_*i*_ }^*n*^_*i*=1_, and model parameters *L, F, µ, τ*, we can compute the likelihood as

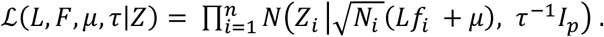

To model our uncertainty in *L, F, µ*, we take a full Bayesian approach in the lower dimension latent space similar to a Bayesian PCA model^16^ as,

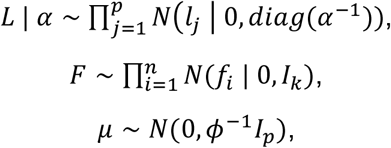

where *α* ∈ *R*^*k*×1^_*>*0_ (*ϕ >* 0) controls the prior precision for variant loadings (intercept). To avoid overfitting, and “shut off” uninformative factors when *k* is misspecified, we use automatic relevance determination (ARD)^16^ and place a prior over *α* as

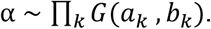

Lastly, we place a prior over the shared residual variance across GWAS studies as *τ ∼ G*(*a*_*τ*_, *b*_*τ*_). We impose broad priors by setting hyperparameters *ϕ* = *a*_*k*_ = *b*_*k*_ = *a*_*τ*_ = *b*_*τ*_ = 10^*−*5^.

### Variational inference

Given our FactorGo model and observed Z-scores summary data, we would like to infer the posterior distribution of parameters *L, F, µ, α, τ*. Unfortunately, there is no closed form expression for learning the posterior exactly, thus, we leverage variational inference to infer an approximate posterior distribution^16,17^. Let *D* be the observed Z-scores and respective GWAS sample sizes. In brief, the true posterior distribution *P*(*L, F, µ, α, τ* | *D*) is approximated by a factorized tractable distribution from the conjugate families

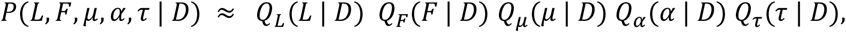

where *Q*_*·*_(*·*) reflects a surrogate approximating posterior for individual model parameters. The optimal functional forms for each *Q* and respective variational parameters are identified by maximizing the evidence lower bound on the marginal likelihood (i.e., ELBO). During inference, variational parameters are updated iteratively until convergence. To further improve the scalability of our approach, we apply a parameter expansion design that converges more rapidly^18^. Namely, after each iteration step, the latent space *F* is centered using a weighted mean and *L* is orthogonalized to reduce coupling effects of latent parameters (see **Supplemental Text 1**). We implemented FactorGo in Python using *Just-In-time* (JIT) compilation through the *JAX* package (see **Web Resources**), which generates and compiles heavily optimized C++ code in real time and operates seamlessly on CPU, GPU or TPU (see **Code Availability**).

### Simulations

To evaluate the performance of FactorGo and tSVD, we performed simulations under a polygenic additive model. Specifically for *i*^*th*^ study, we generated a *p*-vector of true SNP effects *β*_*i*_ as linear combination of *k* latent factors *β*_*i*_ = *Lf*_*i*_, where the values of *L, f*_*i*_ were generated from 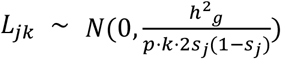 and *f*_*ik*_ ∼ *N*(0, 1), where *j* ∈ [*p*]. The minor allele frequency was sampled from *s*_*j*_ *∼ U*(0.01, 0.5). For simplicity we fixed the intercept *µ* to zeros. Given SNP heritability *h*^2^_*g*_, the total simulated variance in outcome *Y* was *Var*(*Y*_i_) = 1/*h*^2^_*g*_ * Σ_*j*_(*β*_*ij*_ * 2*s*_j_(1 − *s*_*j*_))^2^. Then residuals of each SNP effect in each study became *σ*^2^_*ij*_ = *Var*(*Y*_*i*_) − (*β*_*ij*_ * 2*s*_j_(1 − *s*_*j*_))^2^. Assuming the genotype was centered but not standardized, then the standard errors were 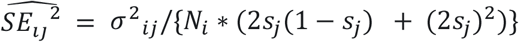 on the per-allele unit, where GWAS sample size *N*_*i*_ was sampled empirically from 2,483 Pan-UK BioBank studies in real data analysis (see **Figure S4**). Finally, we added Gaussian noise to generate observed SNP effects 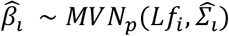 for *i* ∈ [*n*], where the diagonal values of 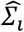 were 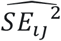. Observed Z-score summary statistics were calculated as 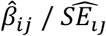.

For each simulated dataset, we applied tSVD and FactorGo on standardized observed Z-score matrices with size *n* × *p* to compare their reconstruction error on true latent parameters. Standardization was applied to columns such that each SNP vector had zero mean and unit variance. Assuming the true model was consistent with FactorGo model and the true number of latent factors *k* was known, we explored extensive scenarios by varying 4 different parameters: number of traits *n* ; 2) number of independent causal SNPs *p* ; 3) number of true latent factors *k* ; 4) additive SNP heritability *h*^2^_*g*_. Each simulated scenario has 30 replications. Next, we examined the influence of model misspecification under four conditions: 1) mis-specified number of latent factors; 2) correlated standard errors due to GWAS sample overlap; 3) no latent factors (i.e., no pleiotropy) and only correlated standard errors; 4) correlated test statistics due to moderate linkage disequilibrium (LD) after LD pruning. Lastly, we examined the robustness of FactorGo across a grid of 5 hyperparameters regarding prior distributions.

### Metrics for simulation

We evaluated the accuracy of FactorGo and tSVD across several metrics. First, to evaluate the accuracy in reconstructed SNP effects matrices *B* = *LF*, we calculated the Frobenius norm between estimates and ground truth, i.e., 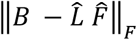. For tSVD decomposition *USV*^*T*^, we defined 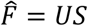 and 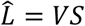. Second, we evaluated the accuracy in estimating variant loadings *L* and factor scores *F*. To account for rotation and scaling in inferred parameters, i.e., 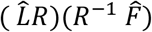 can give the same data likelihood where *RR*^*−*1^ = *I*, we performed procrustes analysis to align the parameters with their ground truth. Briefly, given matrices *A* and *B*, procrustes analysis^19,20^ aims to find a rotation matrix *R* and scaling *s* term such that *min*_*R*_ ||*A − sRB*||^2^_*F*_ subject to *RR*^*−*1^ = *I*. Here, we applied procrustes analysis on the inferred loading matrix 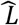 to learn an optimal rotation *R* and scaling factor *s* then computed 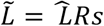, and calculated a final reconstruction error as 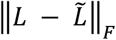. Using the same rotation matrix *R* and scaling factor *s*, we computed 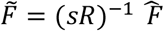 and calculated reconstruction error as 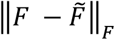.

When no latent factors exist and test statistics correlate across studies due to residual confounding, we applied Levene’s test to compare the variance of inferred parameters. The motivation is that if non-zero error correlation induces false discovery of latent structures, then we expect the variance of 1/*E*(*α*) (or eigenvalues) to deviate from the null of constant variance simulated residual correlations, i.e. 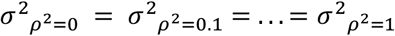.

### Contribution score and squared cosine score

To assess the relative importance of a trait or variant for a given factor, we calculated contribution scores^7^ for FactorGo and tSVD results. Briefly, a contribution score for trait and variant is defined as:

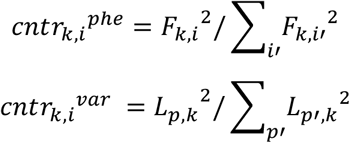

To account for the uncertainty around inferred factors in FactorGo, we first standardized factor scores and variant loadings by their posterior variance before contribution score calculation, i.e. 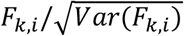 and 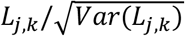. In each pleiotropic factor, we ranked and identified the leading traits or variants based on contribution score (see **Figure 1**). To quantify the relative importance of each factor to a given trait, we calculated squared cosine scores^7^, which are defined as *cos*_*k,i*_^2 *phe*^ = *F*_*k,i*_^2^/Σ_*k*′_ *F*_*k*′,*i*_^2^, where *F* is standardized for FactorGo.

### Quality control on traits from Pan-UK Biobank

Out of the total 7,200 traits from up to 420,531 European individuals in the Pan-UK Biobank (version 04/11/22; see **Web Resources**), we selected traits with number of cases > 1000 for binary traits and total sample size > 1000 for quantitative traits. Pan-UK Biobank ran GWAS using SAIGE to obtain accurate P values for studies with a highly imbalanced ratio of cases to control^21^. For continuous traits, we chose GWAS results under inverse rank normal transformation to correct for outcome distribution. For categorical traits, we selected disease outcomes (see **Table S1**). As a result, the final list consisted of 1,677 binary and 806 quantitative traits (see manifest file in **Table S6**), spanning a wide spectrum of trait domains including diseases, medications, environmental exposures, physical and biomarkers measures, etc. We categorized all 2,483 traits into 9 distinct groups based on the description of UKB field ID (see **Table S1**). We observed marked differences in total sample size across traits, with mean 403,306 for binary traits and 183,577 for quantitative traits (see **Figure S4**).

### Quality control on genetic variants from Pan-UK Biobank

We filtered ∼28 million autosomal variants by INFO score > 0.9, minor allele frequency > 1%, high quality (PASS variant in gnomAD) and high confidence variants (not extremely rare variants) defined by Pan-UK Biobank (see **Figure S5**). Then we excluded the HLA region (Chr6: 25-34Mb on build hg19), indels, and multi-allelic variants. To ensure pleiotropic components across variants, we included SNP variants associated with at least 2 traits using P value threshold 5E-08. Lastly, we applied LD pruning through *Hail* software using the in-sample LD correlation matrix with window size 250 kb and *r*^2^ < 0.3 (see **Web Resources**). These QC steps led to a Z-score data matrix of 51,399 variants by 2,483 traits. 0.002% missing values in Z-scores were imputed using SNP means. For subsequent functional interpretation, we focused only on variants included in the 1000 Genomes Project with functional annotation data^22^ (see **Web Resources**).

### Analyses of Z-score summary data

We implemented both FactorGo and tSVD to learn *k* = 100 latent factors and compare their findings. For FactorGo, we used broad priors by setting all hyperparameters to be 1E-05. For tSVD, we applied the *TruncatedSVD* function from *sklearn* python package with 20 iterations of randomized states (see **Web resources**)^23^. The columns of Z-score data matrix in size *n* × *p* were centered and standardized. The inferred factors were ordered by variance explained in observed data for FactorGo (i.e., *R*^2^) and by singular values for tSVD (see **Supplemental Text 1**). To show robustness of inferred factors subject to choice of *k*, we performed additional analysis using *k* = 90, 110 respectively and compared the top two factors and three leading factors for focal traits in case studies.

### Case studies

To validate results and discover biological insights, we highlighted four traits: BMI and standing height as characteristic polygenic traits, rheumatoid arthritis (RA) as a representative autoimmune disease (a family of diseases known to have substantial shared genetic basis), and prostate cancer (PCa) as the second common cancer for men worldwide with under-explored shared architecture with other traits. For each trait, we characterized the three respective leading pleiotropic factors, and compared results between FactorGo and tSVD.

### Enrichment analysis on variant loadings

To interpret shared biology characterized by inferred factors at the tissue or cell type resolution, we downloaded 205 LDSC-SEG annotations for variants in 1000 Genomes Project^22^ (see **Web Resources**). The annotations are genes specifically expressed in 205 tissue or cell types (e.g., brain vs. non-brain cell types). To leverage the machinery of stratified LD score regression^2,24^ (S-LDSC; see **Web Resources**) for identifying enriched annotation in variant factor loadings, we first transformed the loadings to Z-score scale. To achieve this, we defined a pseudo sample size for each factor as a weighted sum of GWAS sample sizes *N*_*k*_^*pseudo*^ = *∑*_*i*_ *cos*_*k,i*_^2 *phe*^ *· N*_*i*_. Then we created pseudo Z-score by multiplying *N*_*k*_^*pseudo*^ *· L*_*j,k*_ as the Z-score input for S-LDSC software. The LD scores were calculated using n=489 European ancestry individuals from 1000 Genomes with window size 1 cM. Additionally, the LD scores for regression SNPs were calculated separately as the weight for S-LDSC.

We ran S-LDSC on loading-based Z-scores against each annotation to identify enriched tissue or cell type (see **Figure 1**), conditioning on baseline annotations described elsewhere^25^. We used flag *--n-blocks 4000* to obtain a more accurate standard error with 4000 jackknife blocks instead of default 200 since analyzed SNPs were LD-pruned. We calculated q value to control factor-wise FDR < 0.05 using the *qvalue* R package by fixing *λ* = 0, which is equivalent as Benjamini Hochberg adjusted p value^26^. Note that the null distribution of P values from S-LDSC is not uniform because it is a one-sided test for positive coefficient, thus it is not appropriate to estimate the proportion of null hypothesis using the q value method^27^. To demonstrate that our S-LDSC approach is well calibrated, we created 10 non-overlapping annotations for randomly selected gene sets from ∼20,000 genes and computed the enrichment of these annotations over all factors at FDR < 5%. To compute the specificity of enriched tissue or cell types between inferred factors, we calculated all pairwise Jaccard indexes. Briefly, the Jaccard index measures the similarity between two sets *A, B* by 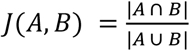, which is the ratio of the number of shared elements over the total number of unique elements.

## Results

### Method evaluation in simulations under model assumptions

We assessed the performance of FactorGo in learning latent parameters across different simulated genetic architectures and compared results with tSVD as a baseline.

First, we found that FactorGo outperformed tSVD exhibiting lower reconstruction error in trait factor scores *F* across all simulated scenarios (Wilcoxon P=3.64E-109; **Figure 2A** and **Figure S1**). Moreover, we observed the FactorGo error in trait factor scores *F* decreased with the increasing number of traits (P=2.09E-24; **Figure S1A**) and number of true latent factors (P=7.30E-26; **Figure S1C**). Error in *F* remained roughly constant across varying numbers of causal SNPs (P=0.99; **Figure S2B**) and average SNP heritability (P=0.36; **Figure S1D**).

**Figure 2.**
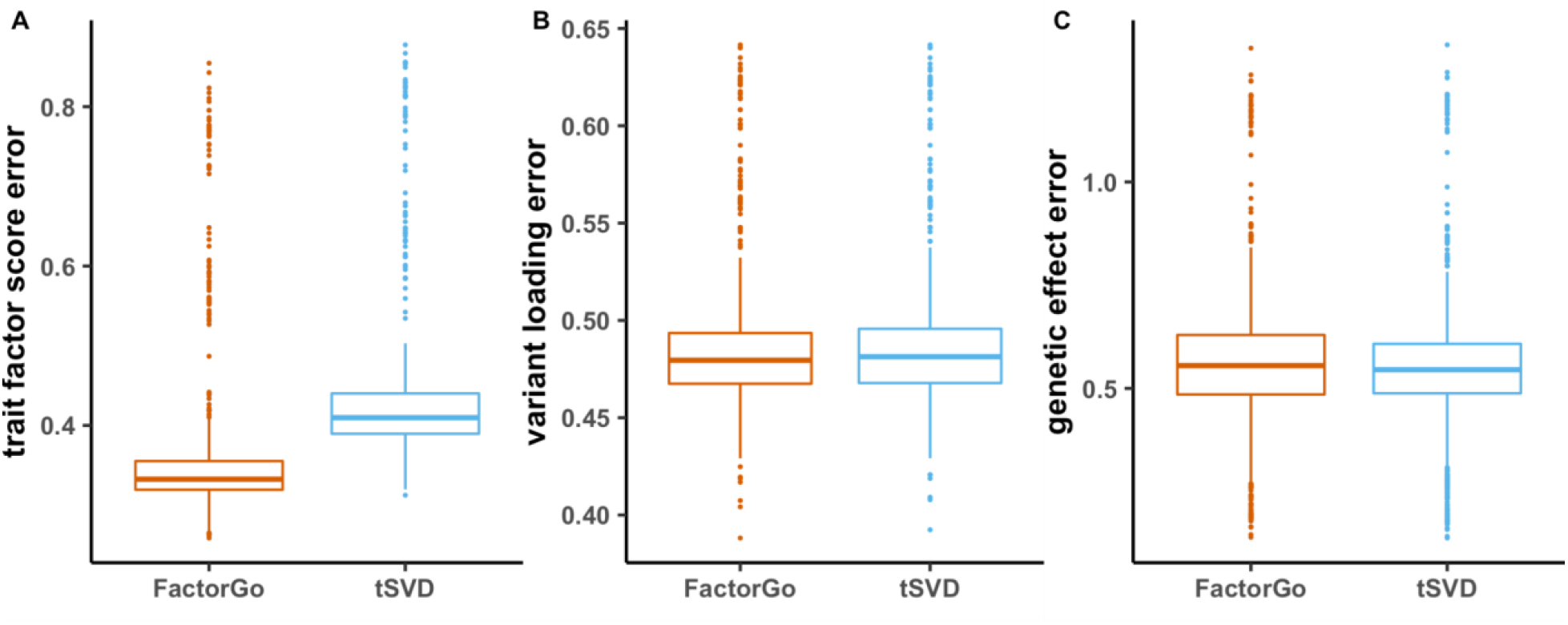
FactorGo provides accurate estimates of model parameters. We report errors for **(A)** trait factor score *F*, **(B)** variant loading *L* and **(C)** genetic effect *B* aggregated over four sets of simulations letting varying either the number of studies (*n*), the number of SNPs (*p*), the number of true latent factors (*k*), and SNP heritability (*h*^2^_*g*_) (See separate results in **Figure S2**). The median value is displayed as a band inside each box. Boxes denote values in the second and third quartiles. The length of each whisker is 1.5 times the interquartile range. All values lying outside the whiskers are considered to be outliers.

Second, although error of variant loading *L* was not significantly different between FactorGo and tSVD (P=0.29; **Figure 2B**), we found FactorGo error decreased with increasing number of traits (P=5.22E-15; **Figure S1A**), number of true latent factors (P=1.40E-23; **Figure S1C**) and average SNP heritability (P=0.071; **Figure S1D**). The error in loadings increased with increasing causal SNPs (P=8.40E-06; **Figure S1B**). The accuracy in genetic effect *B* estimation was not statistically different between FactorGo and tSVD (P=0.10; **Figure 2C**).

Overall, our simulations demonstrate FactorGo provides similar estimates of model parameters as tSVD, with a significant improvement of trait factor scores.

### Method evaluation in simulations under model misspecification

Next, we sought to assess the performance of FactorGo under various settings reflecting model misspecification. First, when the specified *k* differs from the true number of latent factors. When the true number of latent factors *k* = 10, FactorGo performed similarly as tSVD in estimating trait factor scores *F* across varying *k* from 2 to 20 (P=0.21; **Figure 3A**). However, FactorGo provided more accurate estimates in trait factor scores *F* than tSVD (P=0.027) when *k* is underspecified (*k <* 10), compared to when *k* overspecified (*k >* 10; **Figure 3A**). For variant loading *L*, the error was not significantly different between FactorGo and tSVD (P=0.25; **Figure 3B**). Interestingly, the estimates for genetic effects *B* was more accurate in FactorGo (P=0.047) across different *k*, especially when *k* was overestimated (P=2.48E-17; **Figure 3C**).

**Figure 3.**
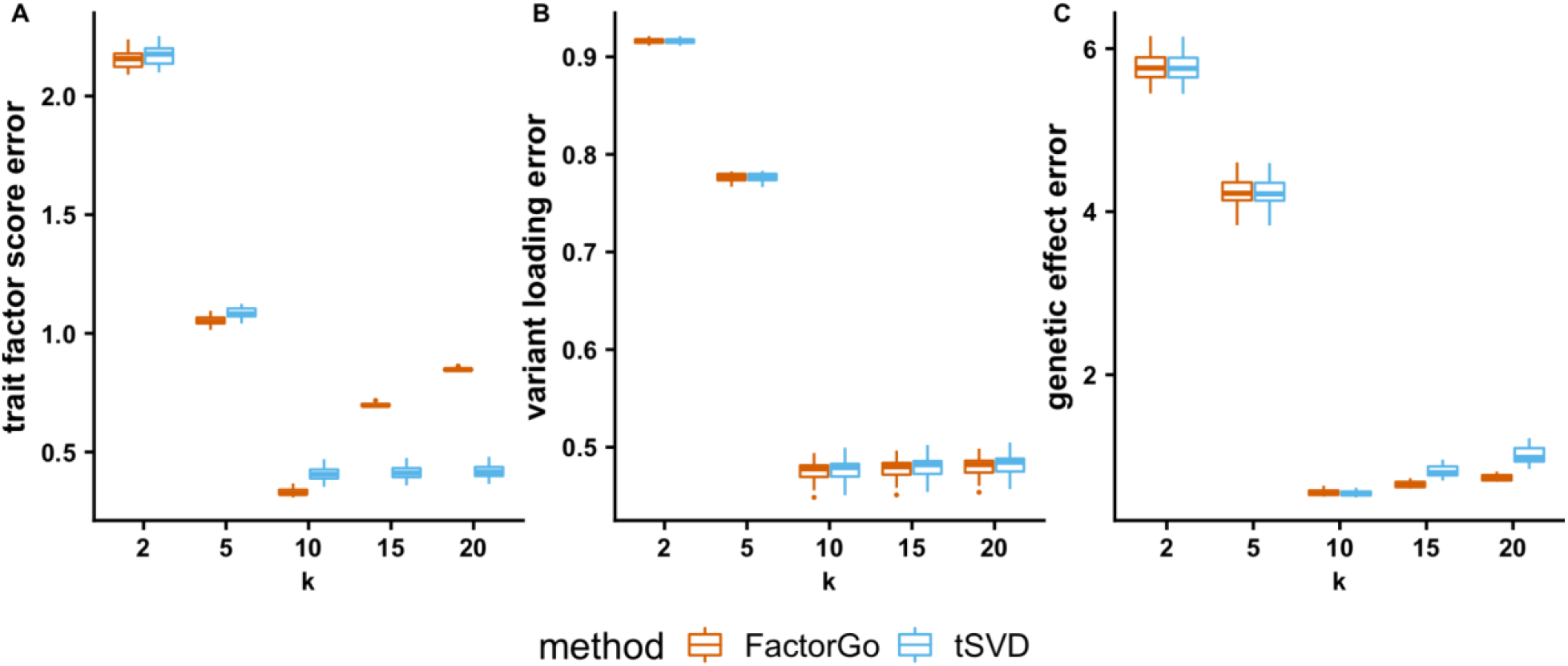
FactorGo outperforms tSVD in trait factor scores when k is underspecified. We report reconstruction error for **(A)** trait factor score *F*, **(B)** variant loading *L*, and **(C)** genetic effect *B* in simulations under varying user-defined latent dimensions *k* = 2,5,10,15,20 when fixing true *k* = 10 (and *p* = 2000, *n* = 100, and *h*^2^_*g*_ = 0.1).

Second, when standard errors and test statistics are correlated due to non-zero LD between SNPs, we observed FactorGo consistently outperformed tSVD in reconstructing trait factor scores (P=2.30E-78; **Figure S2A-C**). FactorGo was especially robust across varying magnitudes of correlated standard errors in estimating trait factor scores (P=1.00) and variant loadings (P=0.90; **Figure S2A**). Third, when no latent factors exist and correlated standard errors across traits due to unmeasured confounding (i.e., shared environment), we found little evidence of latent factor signals in 1/*E*(*α*) from FactorGo (P=1.00) or eigenvalues from tSVD (P=1.00; **Figure S2D**), suggesting both approaches are robust to this confounding.

Lastly, we evaluated the sensitivity of FactorGo to choices of 5 hyperparameters involved with *α* (i.e., prior loading variance), *µ* (i.e., average SNP effect), and *τ* (i.e., residual heterogeneity). For each of the scenarios, we found FactorGo was robust to varying choices of these values in estimating true effects (P=0.96), trait factor scores (P=0.93) and variant loadings (P=0.90; **Figure S3**).

Overall, our simulation results demonstrate that FactorGo accurately identifies latent representation of traits when *k* is underestimated, when test statistics across SNPs are correlated due to LD, and when standard errors are correlated across traits due to unmeasured confounding (i.e., shared environment).

### FactorGo improves interpretation of the pleiotropic components of 2,483 UK Biobank traits

Having demonstrated the performance of FactorGo in simulations, we next characterized 100 pleiotropic factors of 2,483 real traits from the Pan UK Biobank (mean N=331,980; see **Web Resources**). We selected traits by their case or total sample size > 1000. Initial screening on ∼28 million variants by INFO > 0.9 and minor allele frequency > 1% resulted in 8,449,689 high quality common variants. We retained 7,624,608 bi-allelic non-HLA SNP variants and found 1,037,929 of them associated with at least 2 traits at P value < 5E-08. Next, we subsetted to 1,023,655 variants with LDSC-SEG annotation data followed by LD pruning with window size 250 kb and *r*^2^ < 0.3. Finally, we constructed a matrix of GWAS z-scores at 51,399 non-HLA LD-pruned SNP variants across each of the 2,483 traits (see **Methods**). On average each GWAS trait has 109 (SD=541) significant variants. We applied FactorGo and tSVD to the QC’d Z-score matrices to learn 100 pleiotropic factors. Both methods required approximately the same amount of runtime (∼10 minutes for FactorGo on 2 GPUs; **Figure S6**) and explained similar amounts of variance in observed data (38.07% vs. 37.76%). For each method, we ranked factors by the proportion of variance explained.

First, we reported the projection of all traits over the top two FactorGo pleiotropic factors in **Figure 4**. Factor 1 was driven by body weight and basal metabolic rate, and factor 2 was driven by human standing height. We obtained similar patterns for tSVD factors (**Figure S7**). Interestingly, only FactorGo implied the shared comorbidity of COVID-19 with BMI-related traits^28^ in factor 1. Characterization of factors 1 and 2 are given in the section “Characterizing shared biology in FactorGo pleiotropic factors” below. Reminder leading factors were primarily driven by traits with higher heritability compared with factors that explained less z-score variance (P=1.99E-17 and 2.99E-18, respectively; **Figure S8**), which is consistent with heritability reflecting variation in allelic effect sizes.

**Figure 4.**
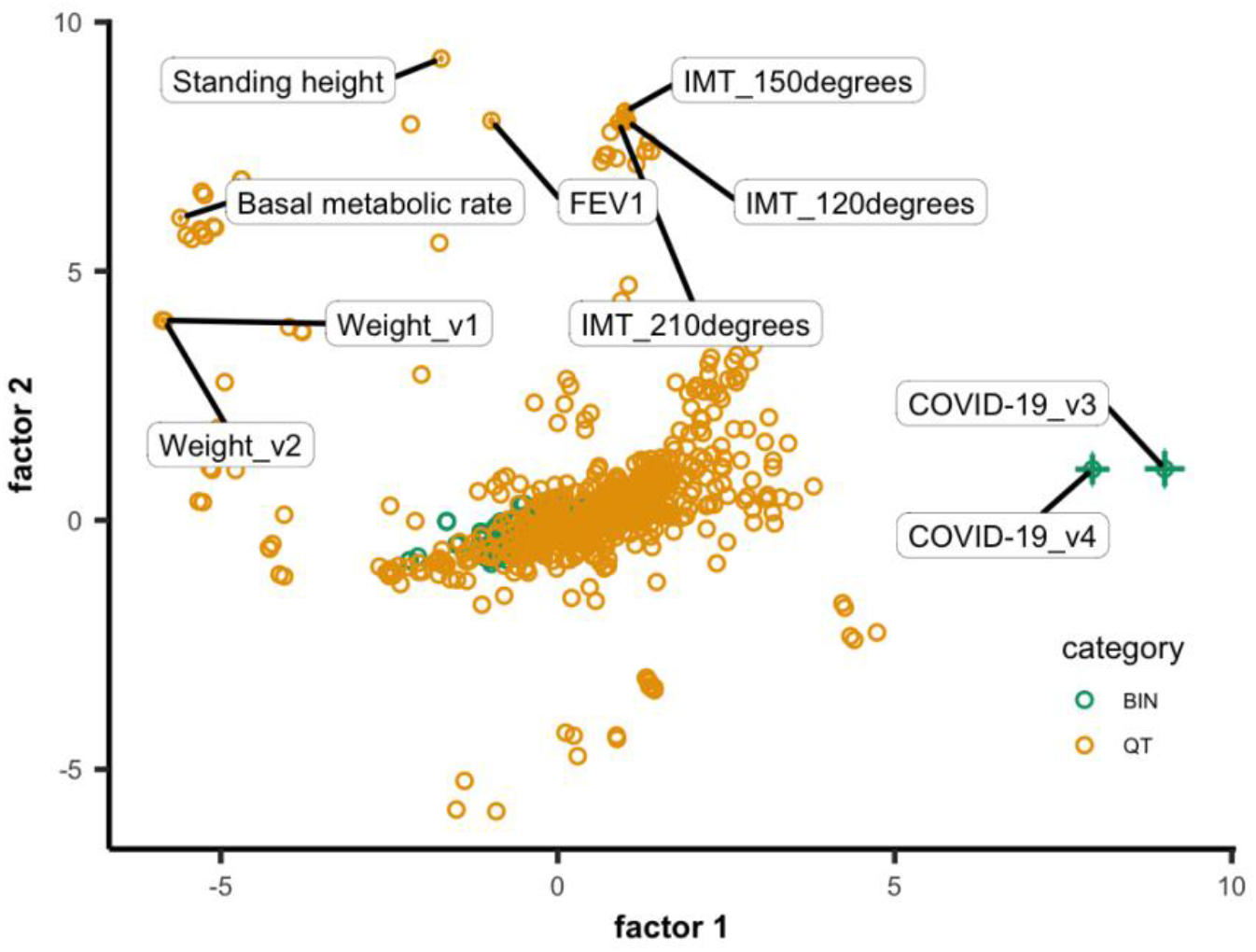
FactorGo factors explain more variance in traits than tSVD factors. We report the projection of 2,483 UK Biobank traits over the top two FactorGo pleiotropic factors. Error bars were 2 times square root of posterior variance for trait factor scores and plotted only for highlighted traits. Binary (BIN) and quantitative (QT) traits were colored differently. FEV1: Forced expiratory volume in 1-second; IMT: Mean carotid intima-medial thickness; “v” for different versions of trait.

Second, by quantifying and ranking the relative importance of pleiotropic factors related to a trait using squared cosine scores (see **Methods**), we observed that the cumulative squared cosine score for each trait was higher in FactorGo than in tSVD at each rank of pleiotropic factor (P<0.05/99; **Figure S9**). To evaluate the sufficiency of these 100 factors in explaining genetic associations from observed data, we found the variance explained by each factor leveled off quickly for both FactorGo and tSVD (**Figure S10A**). The posterior mean of prior precision parameter *α* tracked closely with the variance explained by each factor, suggesting that FactorGo successfully shrunk less informative factors (**Figure S10B**). Finally, to show robustness of FactorGo results with respect to choice of *k*, we performed additional analysis using *k*=90 and 110. The top two latent factors were highly consistent in 20 leading traits and 10 leading variants across *k*=90,100, and 110 results (**Figure S21**).

Third, we evaluated the ability of FactorGo and tSVD to identify relevant shared biology demonstrated by computing tissue-specific enrichment of factor-specific loadings using S-LDSC (see **Methods**; we note that this method was well calibrated under FDR < 5%; **Figure S11**). Overall, we found that the S-LDSC coefficient Z statistics were higher in FactorGo compared with those from tSVD (mean 0.051 vs. -0.042, P=2.58E-10; **Figure S12**). Of the 100 FactorGo factors, we observed 69 were enriched with at least one tissue or cell type at factor-wise FDR < 5%, in contrast to only 40 when using tSVD. FactorGo factors were enriched with 7 tissue or cell types on average and spanned 191/205 tissue or cell types, compared with 130/205 from tSVD (P=6.59E-13). To show specificity of enriched tissue or cell types between inferred factors, we calculated all pairwise Jaccard indexes and found the mean similarity for FactorGo is 0.030, which is lower than 0.045 in tSVD (P=9.37E-04).

Altogether, our results demonstrate that FactorGo identifies biologically meaningful pleiotropic components at the tissue and cell type resolution.

### Characterizing shared biology in FactorGo pleiotropic factors

To characterize the pleiotropic factors identified by FactorGo, we analyzed the leading factors of four representative traits: BMI, height, rheumatoid arthritis (RA), and prostate cancer (PCa). For each trait, we identified its most relevant factor using squared cosine scores, identified the other traits leading this factor using contribution scores, identified the genetic variants leading this factor using contribution scores, and characterized the biology of this factor using S-LDSC on 205 tissue and cell-type-specific annotations (see **Methods**). We assessed that our results were overall consistent across *k*=90,100,110 (**Figure S22-25**).

#### BMI is characterized by Factor 1, associated to brain cell-types

The leading factor for BMI was factor 1 (squared cosine score: 58.85%), which was characterized by body weight (contribution score: 2.32%), basal metabolic rate (2.08%) and body fat masses (cumulative 17.74% across 13 traits; **Figure 5A, Table S2**). The leading variants were proximal to genes such as *WRN* associated with Werner Syndrome (and thus short stature and abnormal fat distribution^29^; rs2553268:G>T: 0.026%) and *TMEM18* associated with obesity (rs13029479:G>A: 0.024%; rs74676797:G>A: 0.024%)^30^. Out of the 33 tissues and cell types significantly enriched in factor 1, 31 were brain cell types including the limbic system and hippocampus (**Figure 5A**), which is consistent with previous findings of brain-specific enrichments in BMI genetic data^2,25^. The next two leading factors for BMI (factor 4 and 7) identified its shared biology with pharynx and digestive tissues respectively (**Supplemental Text 2; Figure S13**). We performed the same analysis using results from tSVD and found no enrichment of cell types in the leading factor for BMI, despite similarly characterized body fat traits (**Figure S17**).

**Figure 5.**
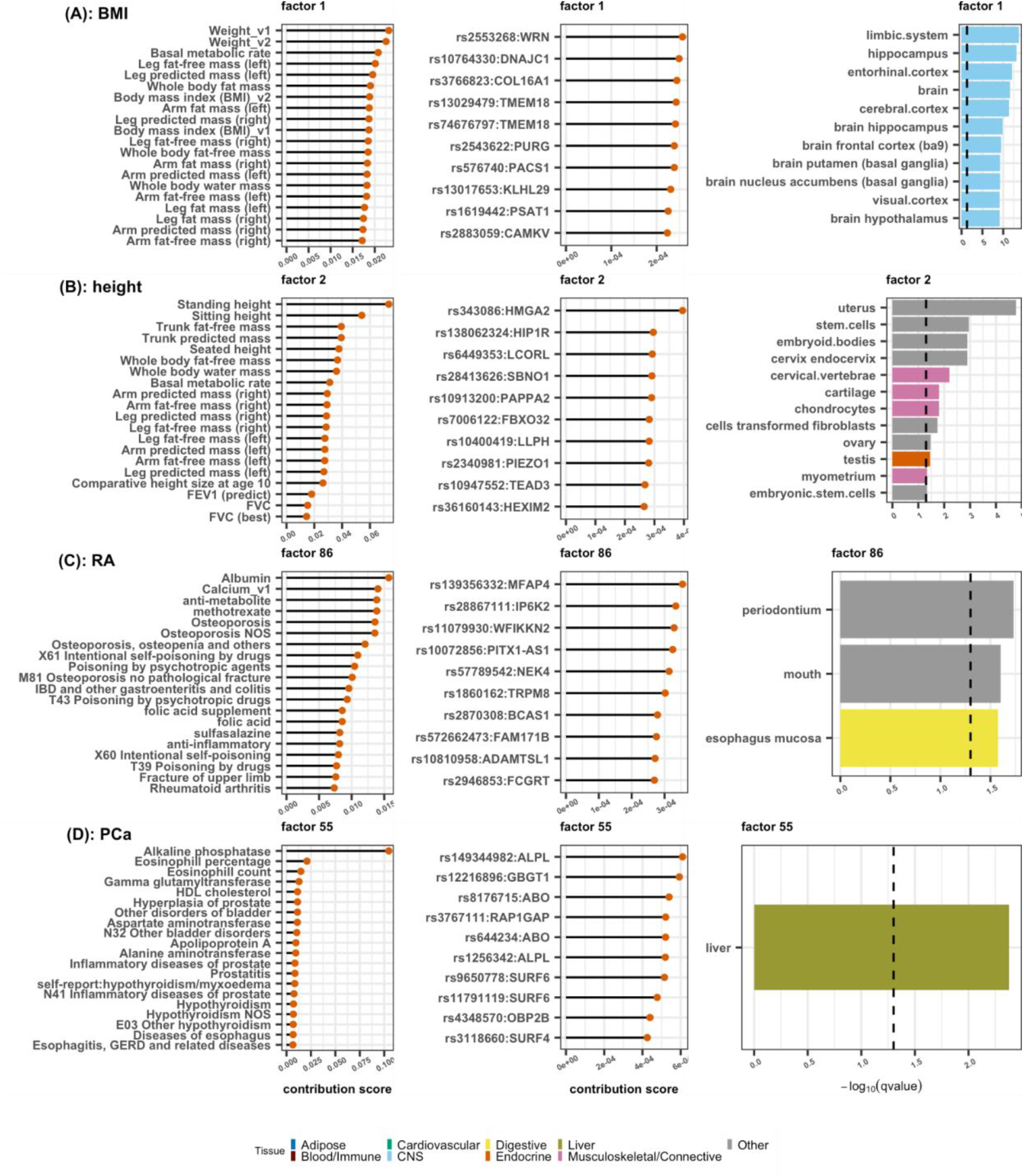
Characterizing shared biology in pleiotropic factors leading four representative traits. We characterized the pleiotropic factors leading **(A)** BMI, **(B)** height, **(C)** rheumatoid arthritis (RA), and **(D)** prostate cancer (PCa). For each focal trait (row), we identified its leading factor, and reported the contribution scores of the 20 leading traits of this factor, the 10 leading variants with their closest gene, and *−log*_10_(*qvalue*) at FDR < 5% for significantly enriched LDSC-SEG annotations (truncated to 10 if more than 20 enriched annotations). See detailed result in **Table S4**. FEV1: Forced expiratory volume in 1-second; FVC: Forced vital capacity; “v” for different versions.

#### Standing height is characterized by Factor 2, associated with musculoskeletal tissues

As the leading factor for standing height, factor 2 (squared cosine score: 38.67%) characterized leading traits as standing height (7.36%), sitting height (5.41%), and body fat masses (1.39%; **Table S2**). These associations were driven primarily by an intron variant in height-associated gene *HMGA2* (rs343086:T>C: 0.04%)^31,32^. As expected, factor 2 exhibited enrichment for musculoskeletal tissues such as cartilage and chondrocytes (**Figure 5B**). Additionally, we replicated enrichment for reproductive organs such as uterus and cervix^25,33^. This result is also consistent with prior work demonstrating that overexpression of *HMGA2* alters production of growth hormone in mice^34^, in addition to reproductive tissue development^35^. The next two leading factors for height suggested a shared biology with cardiovascular and immunity, respectively (**Supplemental Text 2; Figure S14**). For tSVD, we found its leading factor similarly characterized height traits, but did not exhibit evidence of cell-type enrichment (**Figure S18**).

#### Rheumatoid arthritis (RA) leading factor is driven by inflammatory mechanisms

For RA, factor 86 (squared cosine score: 7.17%) was explained primarily by inflammation-related traits (**Figure 5C**), such as blood albumin level (1.57%), blood calcium level (1.40%), methotrexate (a common treatment for RA; 1.39%), osteoporosis conditions (cumulative 5.52% across 5 traits; **Table S3**), and other autoimmune diseases such as inflammatory bowel disease (0.96%)^36–38^. We found these signals were driven by variants proximal to genes *MFAP4* (rs139356332:G>C: 0.036%) and *IP6K2* (rs28867111:G>A: 0.033%), both of which are involved with inflammatory mechanisms^39,40^. Interestingly, we observed factor 86 exhibited enrichment in periodontium and mouth (**Figure 5C**), which is supported by prior epidemiological evidence of common periodontal conditions in individuals with RA due to autoantibodies and arthritis triggered by oral pathogens^41^. The next two leading factors for RA (factor 75 and 76) suggested a shared biology with kidney, liver and central nervous system (**Supplemental Text 2; Figure S15**). Different from FactorGo, the leading factor for RA from tSVD characterized IGF-1 measure and cardiac disorders, but not enriched with any cell types (**Figure S19**).

#### Prostate cancer leading factor identifies ALP as a PCa candidate biomarker

For prostate cancer (PCa), the leading factor was factor 55 (squared cosine score: 17.94%), characterized by diseases in prostates, including hyperplasia of prostates (1.13%) and inflammatory diseases in prostates (1.63%) (**Figure 5D**). The leading trait was alkaline phosphatase (ALP) level in blood (10.47%), associated to the leading missense variant in the *ALPL* gene that encodes ALP proteins (rs149344982: G>A: 0.061%). Since ALP is an enzyme mostly produced by the liver and bone, this factor was indeed enriched with genes specifically expressed in the liver. Previous work found higher serum ALP was associated with poor overall survival rate of patients with PCa, which likely reflects bone metastatic tumor load^42^. The next two leading factors for PCa (factor 1 and 58) suggested shared comorbidities of PCa involved with BMI and hormonal disorders (**Supplemental Text 2; Figure S16**), which is consistent with previous works investigating dietary risk factors^43^ as well as the well-documented role of hormonal dependency due to expression of androgen receptor (*AR*) ^44^. Different from FactorGo, the leading factor for PCa from tSVD prioritized corneal resistance factors, geographic home locations and heel bones measures (**Figure S20**). Additionally, tSVD results displayed enrichment for genes expressed specifically in colon, suggesting alternative shared biological mechanisms compared with FactorGo.

## Discussion

In this work, we presented FactorGo to identify and characterize pleiotropic components across thousands of human complex traits and diseases using Z-score summary statistics. Our method enables investigating the phenome-wide shared genetic components while appropriately modeling uncertainty in variant effect estimates. When applied to 2,483 phenotypes from the UK BioBank individuals, we found that FactorGo factors explained more variance on average and were more powerful in identifying shared biology compared with tSVD factors. We validated brain-specific enrichment for BMI factors, muscular skeletal and reproduction enrichment for height factors. For disease traits, FactorGo suggests a shared etiology between rheumatoid arthritis and periodontal conditions. Moreover, we found alkaline phosphatase as a candidate but less established biomarker for prostate cancer, which provided evidence for further experimental validation.

FactorGo has several advantages compared to the scalable but model-free approach tSVD. First, FactorGo learns pleiotropic factors at similar computational cost by leveraging state-of-art variational inference and fast python implementation. Second, we showed using simulations that FactorGo outperformed tSVD in estimating trait factor score under model assumption and model misspecification such as correlated standard errors due to GWAS sample size. Third, in real data analyses, we found more enrichment of tissue or cell types in FactorGo factors than in tSVD factors.

Our tool has several implications for downstream analyses. First, we demonstrated that analyzing phenome wide GWAS summary statistics from biobanks can not only recapitulate known shared biology for traits such as BMI, height and RA, but also nominate candidate biomarkers in diseases for further clinical evaluation such as ALP for prostate cancer. This testifies the benefit of enabling scalability of model-based statistical approaches jointly analyzing thousands of GWAS summary data from large biobanks. Second, leveraging factor loadings within enrichment analysis using differentially expressed gene annotations allowed us to interpret the biology of a given factor at tissue or cell type level. Our implementation of S-LDSC readily allows analyzing other functional annotations such as chromatin accessibility and transcriptional factors.

Although FactorGo has provided robustness in simulations and rich insights in the analyses of UK Biobank phenotypes, it has some limitations. First, our method focused on learning pleiotropic factors from linear genetic effects and ignored non-linear or epistatic effects. While many lines of evidence pointed to linear models capturing the bulk of trait heritability^45,46^, our results also illustrated rich meaningful biological insight that could be obtained from linear effects alone. Second, our model assumes independence of residual errors, which was unlikely to be true given overlapped samples in large biobank GWAS studies. However, we showed in simulation that the estimation of latent parameters was robust to error correlation. Third, FactorGo didn’t outcompete tSVD in estimating variant loadings in our simulations. However, we provided a probabilistic model to account for heterogeneity in summary statistics across GWAS studies without adding extra run-time cost. Fourth, while our method requires to predefine the number of latent factors *k*, our simulations have shown that results are biased if *k* was fixed to a too high value. However, to ensure that this limitation is unlikely to impact our results, we performed additional analysis using *k*=90 and 110. The top two latent factors were highly consistent in 20 leading traits and 10 leading variants across *k*=90,100, and 110 results (**Figure S21**). The leading factors for BMI, height, RA, and PCa were overall consistent in traits (**Figure S22-25**). Fifth, in real data analysis, our selection of variants using genome-wide significance thresholds can underestimate the degree of pleiotropy due to lack of power, especially in disease traits. For example, in the case study of prostate cancer, we did not observe prostate cancer in the top rank of leading factor, suggesting either prostate cancer has limited shared components with other traits or lack of power in GWAS study to estimate the variant effects. Despite this, we were still able to recapitulate known shared biology for BMI, height and RA using this subset of pleiotropic variants. Similarly, our selection of variants involved an LD pruning procedure. While pruning could limit the functional interpretation of the latent factors, our gene-set analyses leveraging LD scores computed on a sequenced reference panel mitigates this issue. We anticipate that improvement in fine-mapping techniques and ongoing efforts to perform fine-mapping on hundreds of phenotypes at the biobank scale^47^ should improve variant selection in the near future. Sixth, unlike other methods based on non-negative matrix factorization^4^, our model did not distinguish between varying directional effects of pleiotropic factors, but rather focused on non-directional summary of pleiotropic effects. Seventh, recent works have highlighted that shared effect sizes across traits might be driven by assortative mating^48^. Further investigation is required to see how it impacts the interpretation of our results. Lastly, although our method was developed for single ancestry analysis, it can be extended to multi-ancestry data and learn shared genetic components. Taking a step further regarding the model and subsequent interpretation, it is also possible to incorporate functional annotation as priors so that interpreting functional enrichment *a posteriori* is more straightforward.

In conclusion, FactorGo provides a variational Bayesian factor analysis model on GWAS summary statistics to learn and characterize pleiotropic factors across thousands of human complex traits and diseases. It allows rich biological interpretation at tissue or cell type specific level.

## Data Availability

This study relied on publicly available summary statistics. All results produced in this work are available online at zenodo: https://doi.org/10.5281/zenodo.7765048

https://pan.ukbb.broadinstitute.org/

## Acknowledgements

This work was funded in part by the National Institutes of Health (NIH) under awards P01CA196569, R01HG012133, R01CA258808, R35GM147789 and R00HG010160.

We thank Dr. Tiffany Amariuta for her comments and suggestions to this manuscript.

## Author Contributions

Z.Z, S.G, and N.M developed the method. Z.Z performed analysis. J.J and A.K prepared data and performed analyses. All authors edited and approved of the manuscript.

## Web Resources

JAX: https://github.com/google/jax

Hail: https://hail.is/docs/0.2/

1000Genome annotations: https://alkesgroup.broadinstitute.org/LDSCORE/

LDSC-SEG annotations: https://alkesgroup.broadinstitute.org/LDSCORE/LDSC_SEG_ldscores/

Pan-UK BioBank: https://pan.ukbb.broadinstitute.org/

## Data Availability

Summary statistics analyzed in this work and results were available on Zenodo^49^: https://zenodo.org/record/7765048#.ZB3KHezMLCA

GWAS summary statistics from Pan-UK BioBank on AWS cloud storage: https://pan-ukb-us-east-1.s3.amazonaws.com/sumstats_flat_files/*

In-sample LD correlation matrix for Europeans released by Pan-UK Biobank: s3a://pan-ukb-us-east-1/ld_release/UKBB.EUR.ldadj.bm

## Code Availability

FactorGo software: https://github.com/mancusolab/FactorGo

FactorGo analysis code: https://github.com/mancusolab/FactorGo_analysis

