## Supplemental figures and tables for "A scalable variational approach to characterize pleiotropic components across thousands of human diseases and complex traits using GWAS summary statistics"

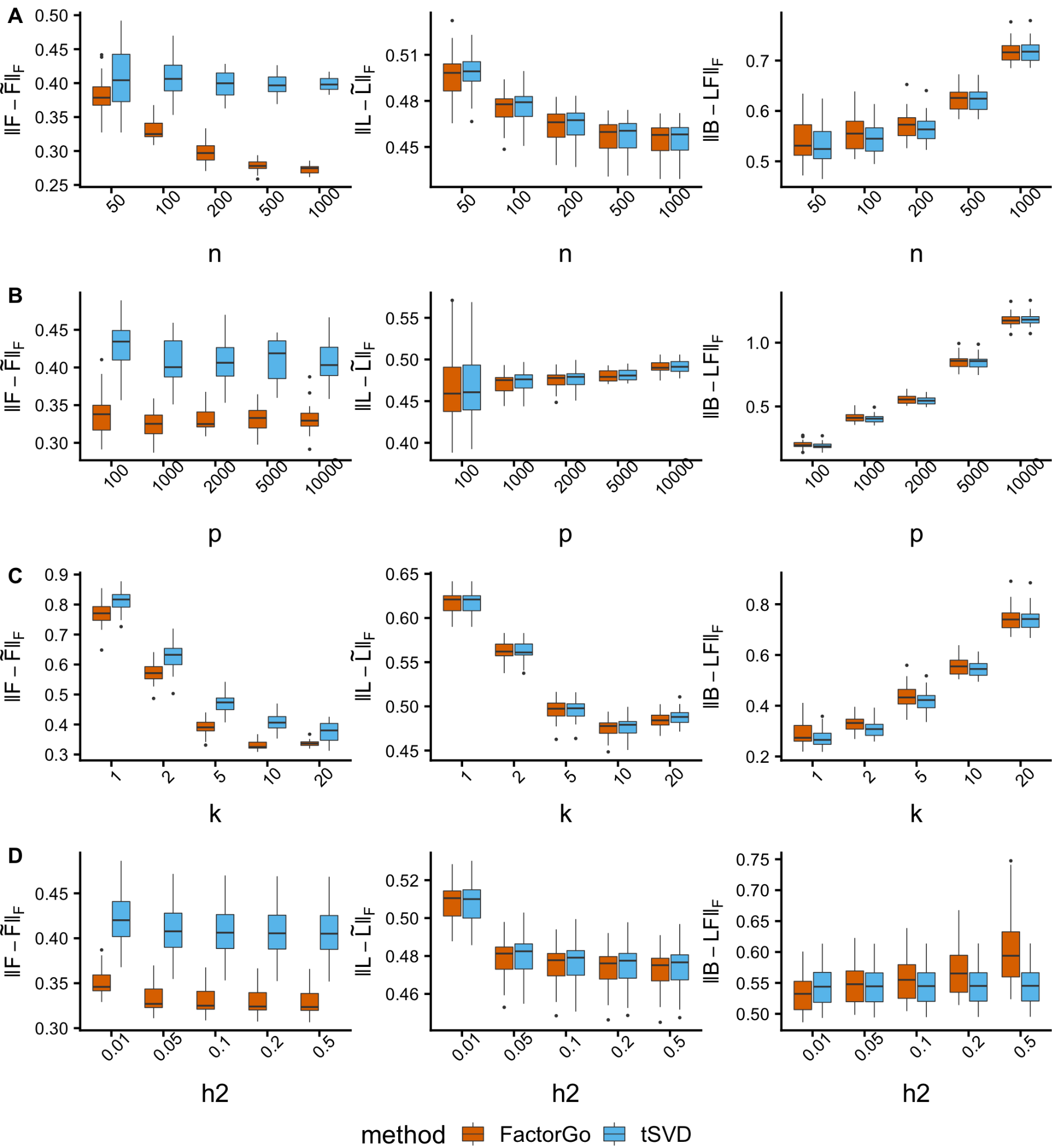

**Figure S1. FactorGo outperforms tSVD in trait factor scores in each scenario under model assumptions.**

Trait factor score error $||F-\tilde{F} ||_{F}$ , variant loading error $||L-\tilde{L} ||_{F}$ and Genetic effect error $||B-LF||_{F}$ under 4 varying parameters: **(A)** number of studies ($n$), fix $p=2000$, $k=10$, ${h^{2}}_{g}=0.1$; **(B)** number of SNPs ($p$), fix $p=2000$, $n=100$, ${h^{2}}_{g}=0.1$; **(C)** number of true latent factors ($k$), fix $p=2000$, $n=100$, ${h^{2}}_{g}=0.1$; **(D)** SNP heritability (${h^{2}}_{g}$), fix $p=2000$, $n=100$, $k=10$.

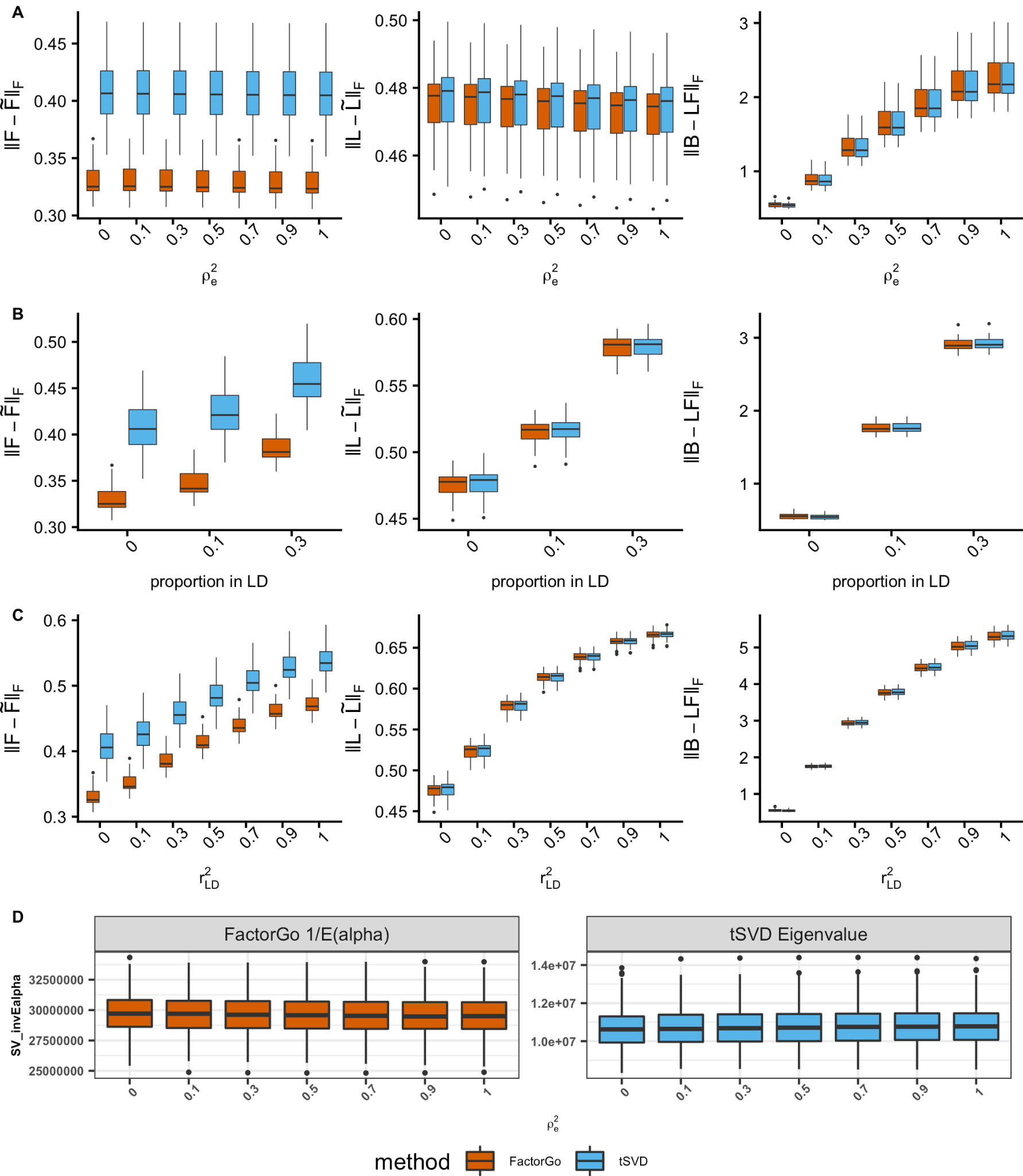

**Figure S2. FactorGo is robust to correlated standard errors and outperforms tSVD in learning trait factor scores.**

Fix $p=2000$, $n=100$, $k=10$, ${h^{2}}_{g}=0.1$, we simulated **(A)** correlated standard errors with varying residual correlation coefficient ${\rho^{2}}_{e}$; adjacent pairs of SNPs in LD with **(B)** varying proportion of SNPs in LD at fixed ${r^{2}}_{LD}=0.3$ or **(C)** different magnitude of correlation at fixed proportion 30%. When there are no latent structures (i.e., no pleiotropy) and only correlated errors **(D)**, we ran both methods with $k=10$ and plotted the $1/E(\alpha)$ from FactorGo and eigenvalues from tSVD methods by correlation magnitude ${\rho^{2}}_{e}$.

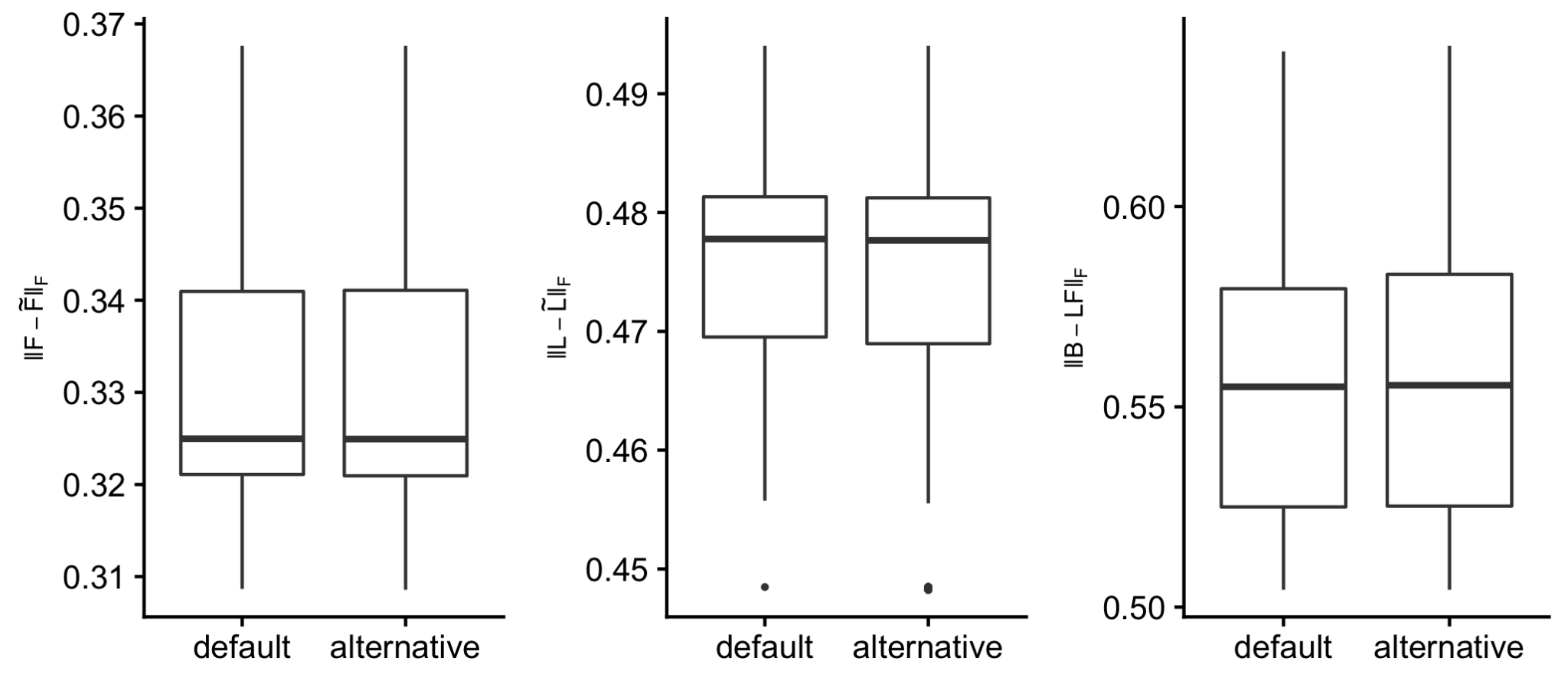

**Figure S3. FactorGo is robust to different choices of hyperparameters in simulations.**

Trait factor score error $||F-\tilde{F} ||_{F}$ , variant loading error $||L-\tilde{L} ||_{F}$ and Genetic effect error $||B-LF||_{F}$ by hyperparameter specification. For each of the 30 simulated data when fixing $p=2000$, $n=100$, $k=10$, ${h^{2}}_{g}=0.1$, we ran FactorGo using all combinations of 1E-05 and 1E-03 for five hyperparameters (total 2^5). The “default” is 1E-05 for all five hyperparameters. Here we compared the reconstruction errors under default setting versus all other alternative settings.

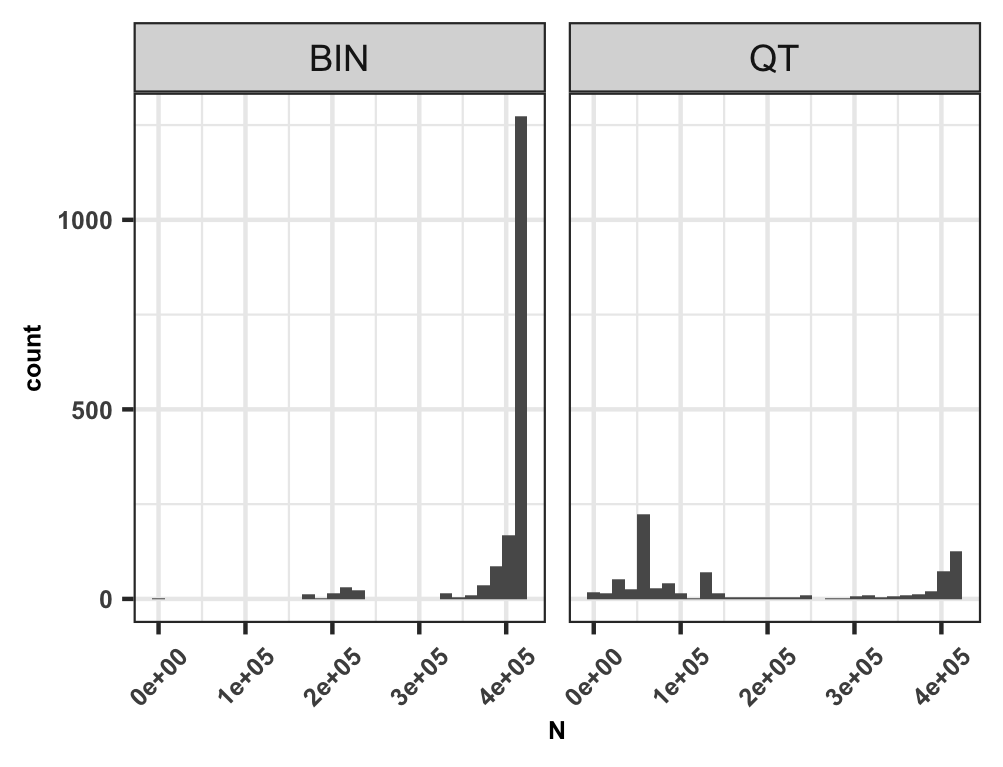

**Figure S4. GWAS sample size distribution in Pan-UK Biobank.**

Histogram of GWAS total sample size of 2,483 studies from Pan-UK Biobank Europeans (max N=420,531) by 1,677 binary (BIN) traits and 806 quantitative (QT) traits. In simulation, the GWAS sample size was sampled empirically from this distribution.

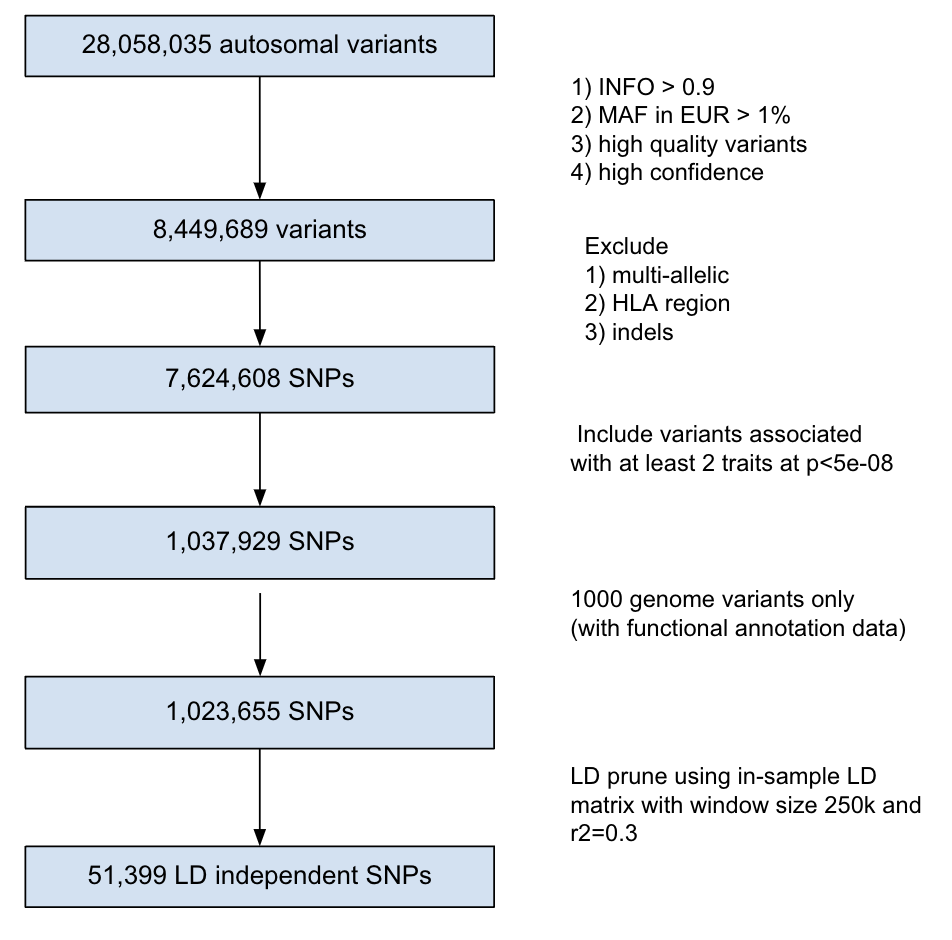

**Figure S5. Summary of genetic variants filtering in real data analysis of Pan-UK Biobank.**

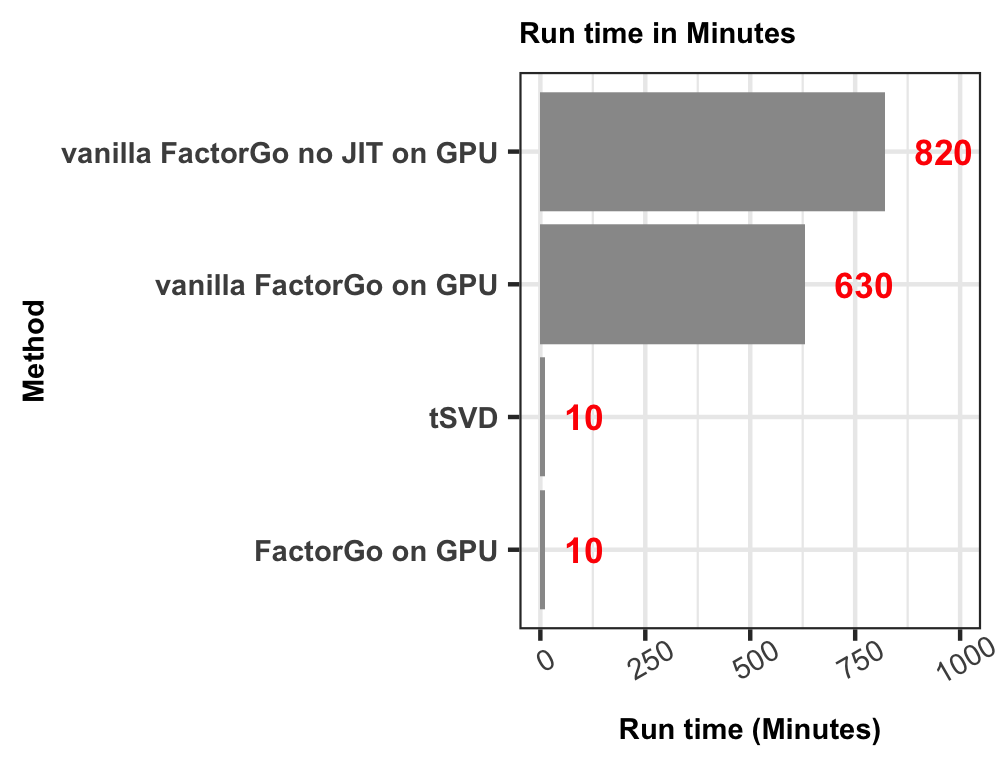

**Figure S6. FactorGo and tSVD have the same run time for real data analysis of 2,483 traits and 51,399 variants.**

In contrast to vanilla FactorGo, the default FactorGo implements a parameter expansion design (**Supplemental Text**). JIT (Just-In-Time) is a fast execution of python code through the *JAX* package (**Web resources**).

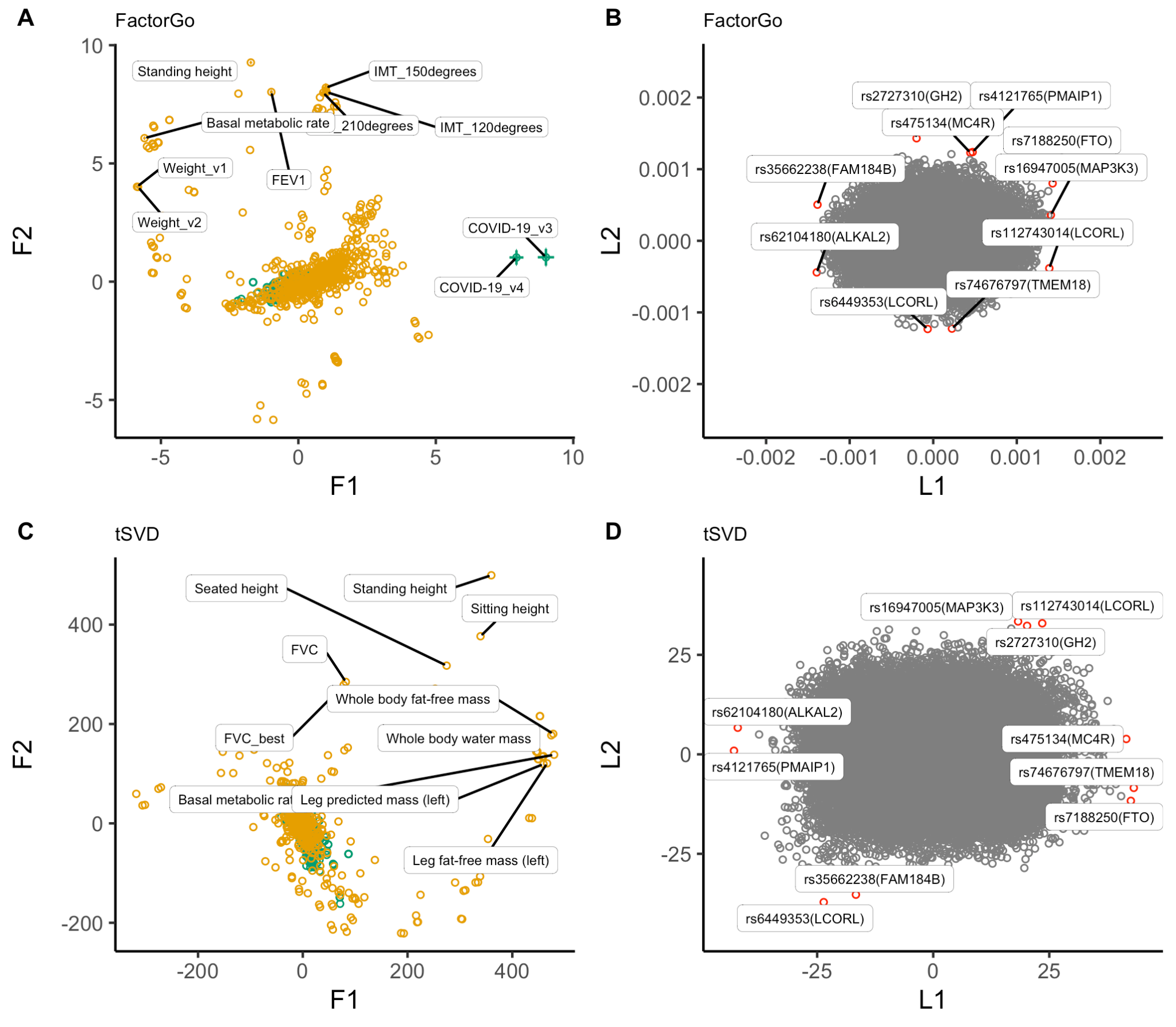

**Figure S7. Trait factor scores and variant loading scores for the top two factors in FactorGo and tSVD.**

We highlighted 10 leading traits and 10 leading variants (red) for top two factors from FactorGo and tSVD. F1 and F2 are factor scores. L1 and L2 are variant loadings. Binary and quantitative traits are colored differently. FEV1: Forced expiratory volume in 1-second; IMT: Mean carotid IMT (intima-medial thickness); FVC_best: Forced vital capacity, best measure.

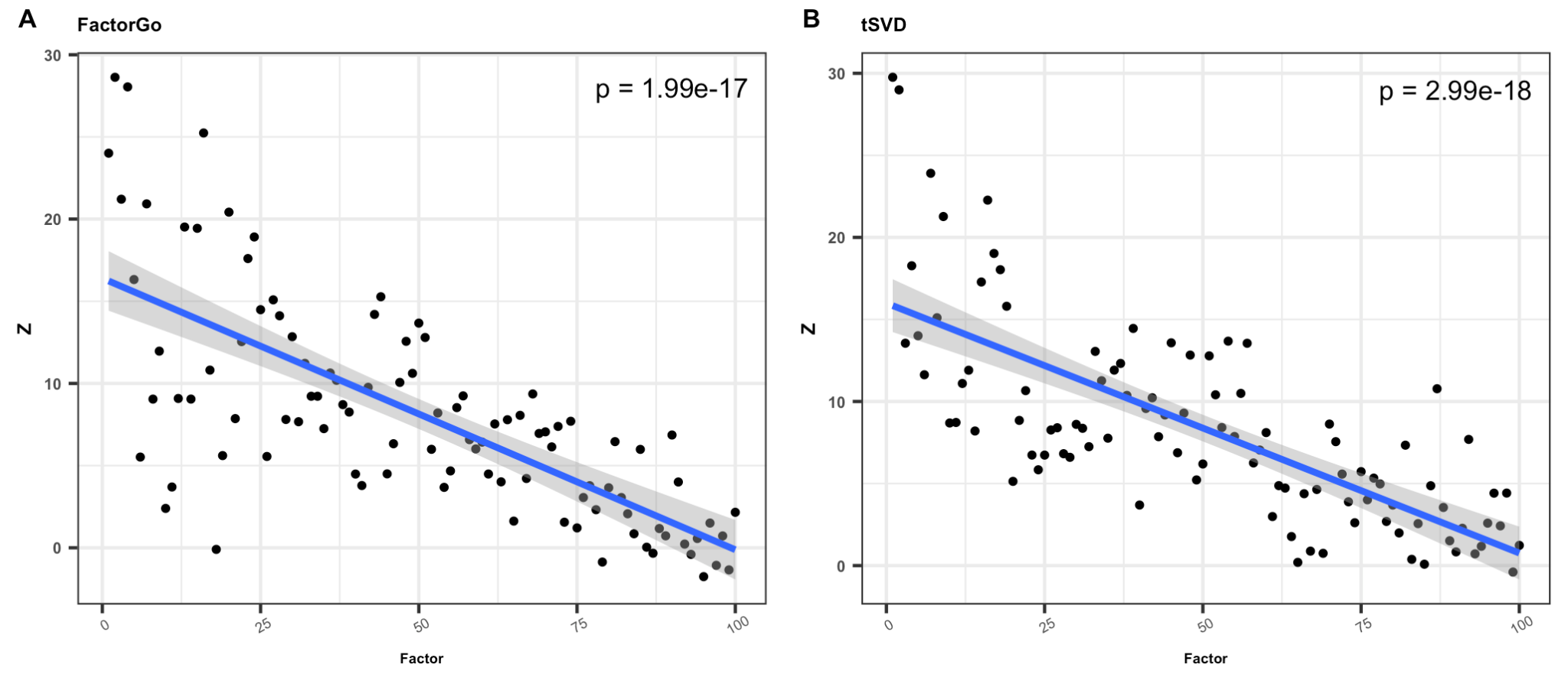

**Figure S8. Factor scores in top factors are driven by traits with high heritability.**

For **(A)** FactorGo and **(B)** tSVD respectively, each point is the test statistics Z-score for linear association between trait factor scores and the observed heritability estimated by LDSC for 2,305 traits with heritability estimates. Blue line is the fitted regression line with gray confidence band over these 100 points on each plot. This association decay with factor rank.

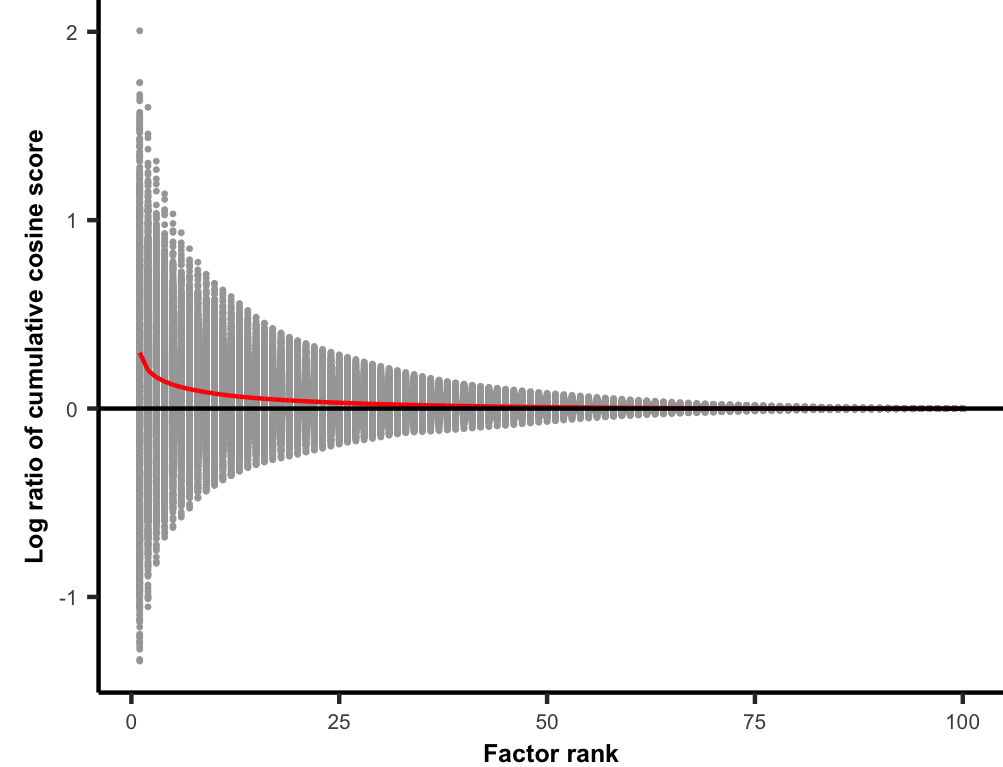

**Figure S9.** **Cumulative squared cosine score for each trait was higher in FactorGo than in tSVD at each rank of pleiotropic factor.**

We report the ratio of cumulative squared cosine score of FactorGo versus tSVD on log scale at each factor rank for each trait. The squared cosine score sums to 1 for each trait so that this ratio approaches log1=0 as rank increases. Red line is the mean of the ratio at each rank across traits.

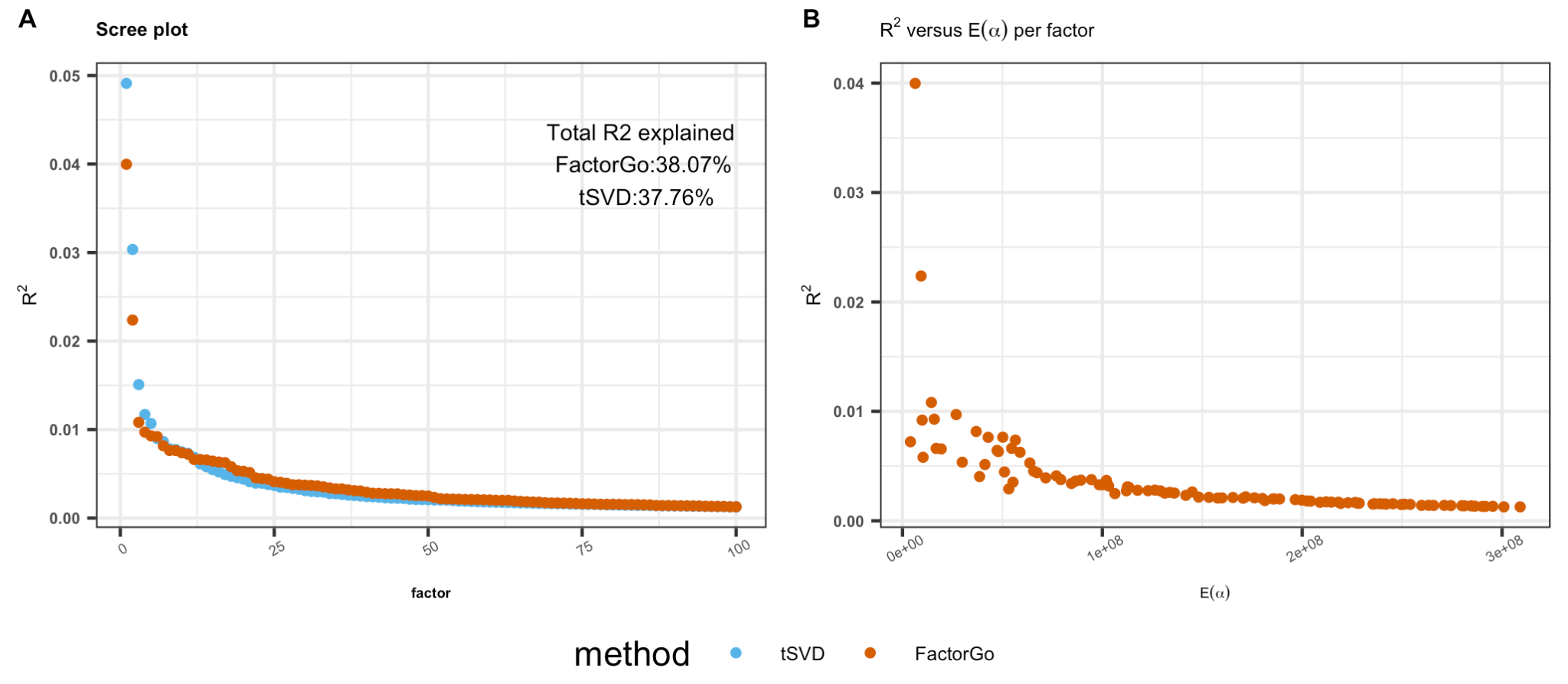

**Figure S10. Variance explained by FactorGo factors tracks closely with posterior mean of ARD parameters** $\alpha$**.**

**(A)** Variance explained ($R^{2}$) by each factor from FactorGo and tSVD. The factors are ordered by $R^{2}$ for both methods. **(B)** Variance explained ($R^{2}$) by each factor in FactorGo versus their posterior mean of ARD prior parameter $\alpha$.

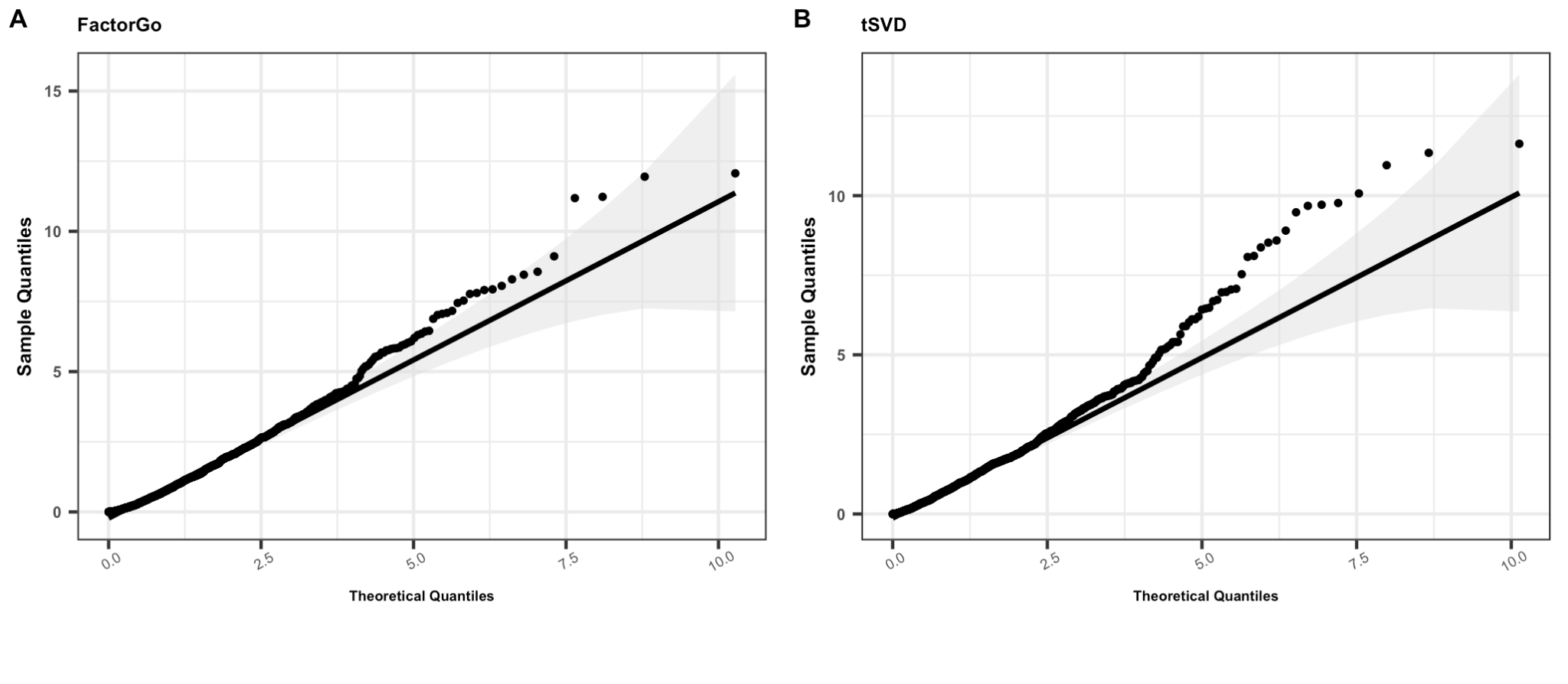

**Figure S11. QQ plot of enrichment P values for randomly selected gene set annotations.**

To show our implementation of S-LDSC is well-calibrated for both **(A)** FactorGo and **(B)** tSVD, we compared P values distribution from S-LDSC enrichment results of 10 randomly selected gene sets to theoretical quantiles. Gray regions are pointwise confidence bands. Given the randomly selected gene sets of size ~2000 genes may be not truly “null”, there is some deviation from null distribution.

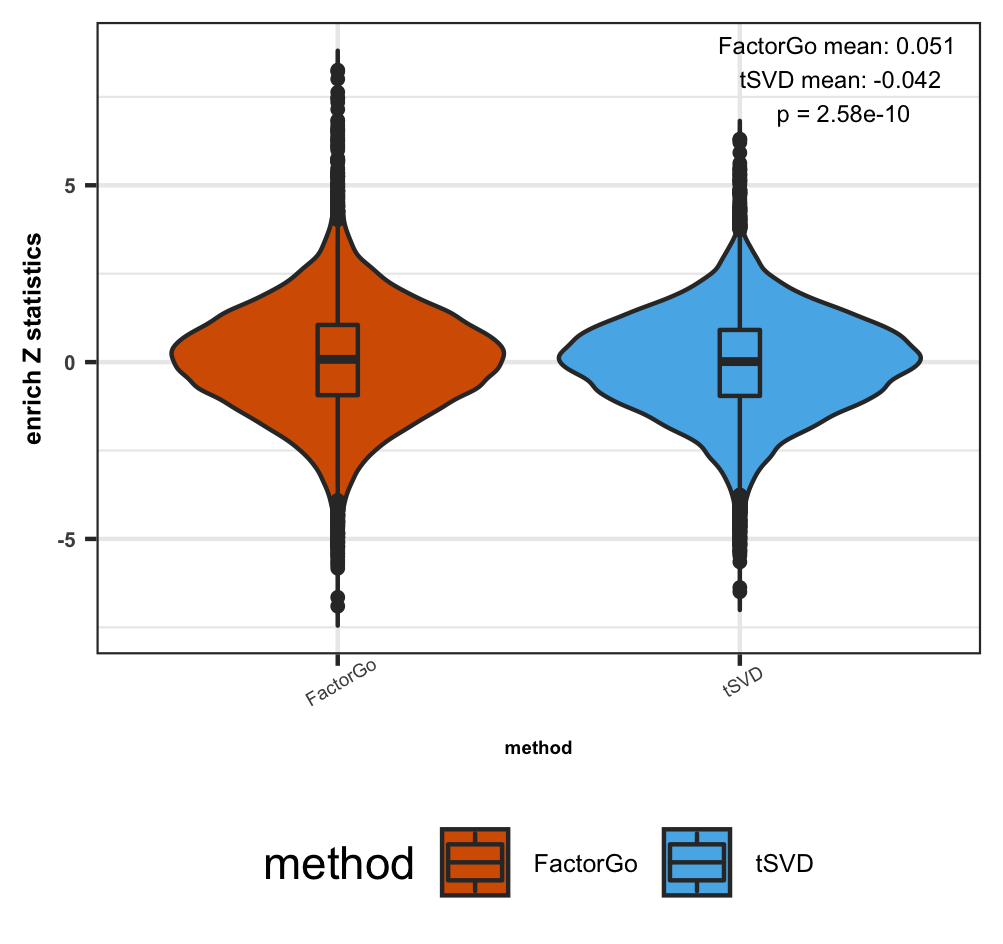

**Figure S12. FactorGo factors show higher enrichment Z test statistics than tSVD.**

Violin plot and embedded boxplot of enrichment Z test statistics (one-sided test) from S-LDSC results for 205 annotations across all 100 factors for each method.

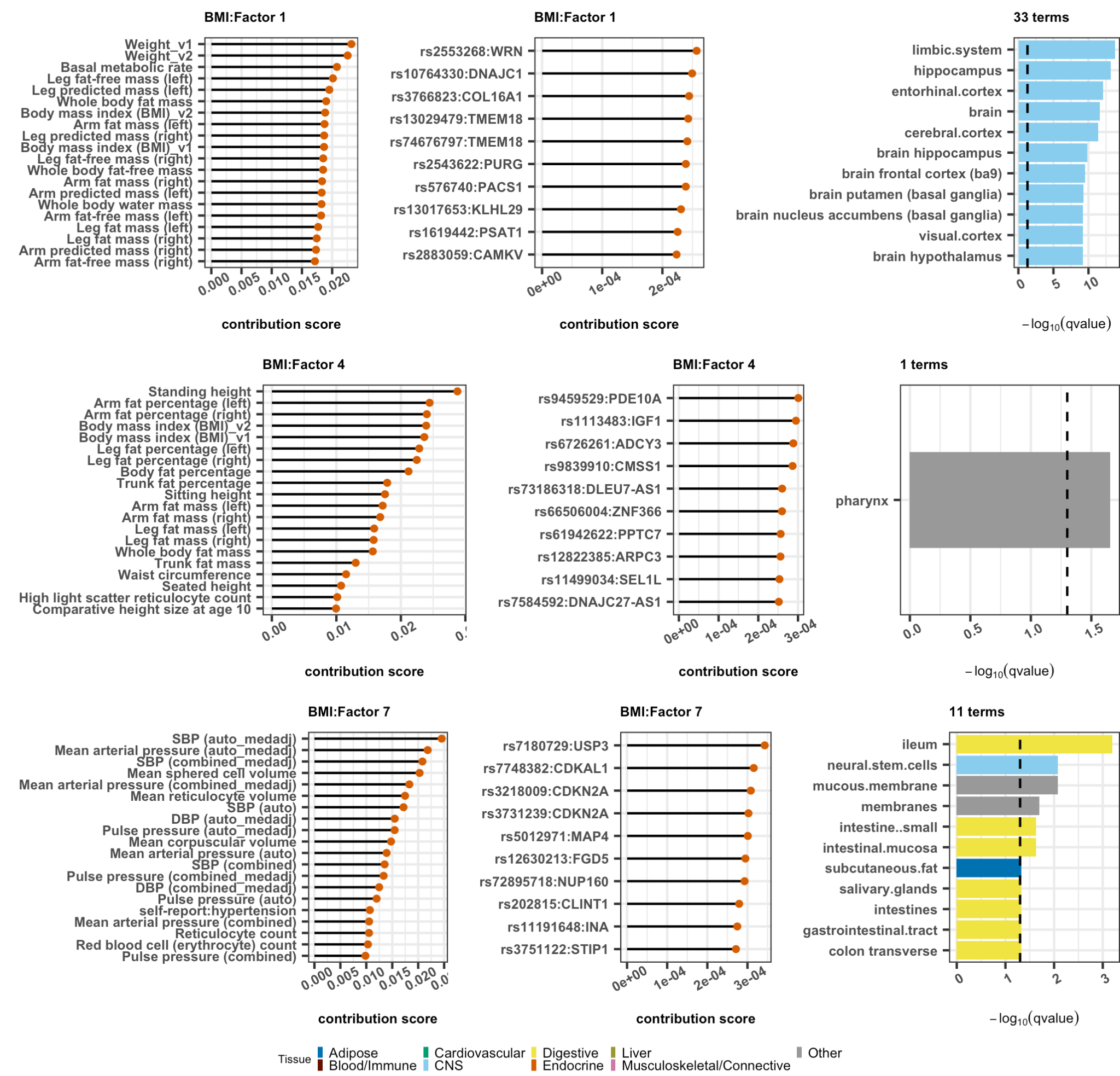

**Figure S13. Characterizing three leading factors in FactorGo for BMI.**

Results for factor 1, 4 and 7 (row) include 20 leading traits, 10 leading variants with closest gene, and enriched LDSC-SEG tissue or cell type (truncated to 10 if more than 20 enriched annotations). Total number enriched annotations are included in the title on the third column. Detailed results in **Table S4**.

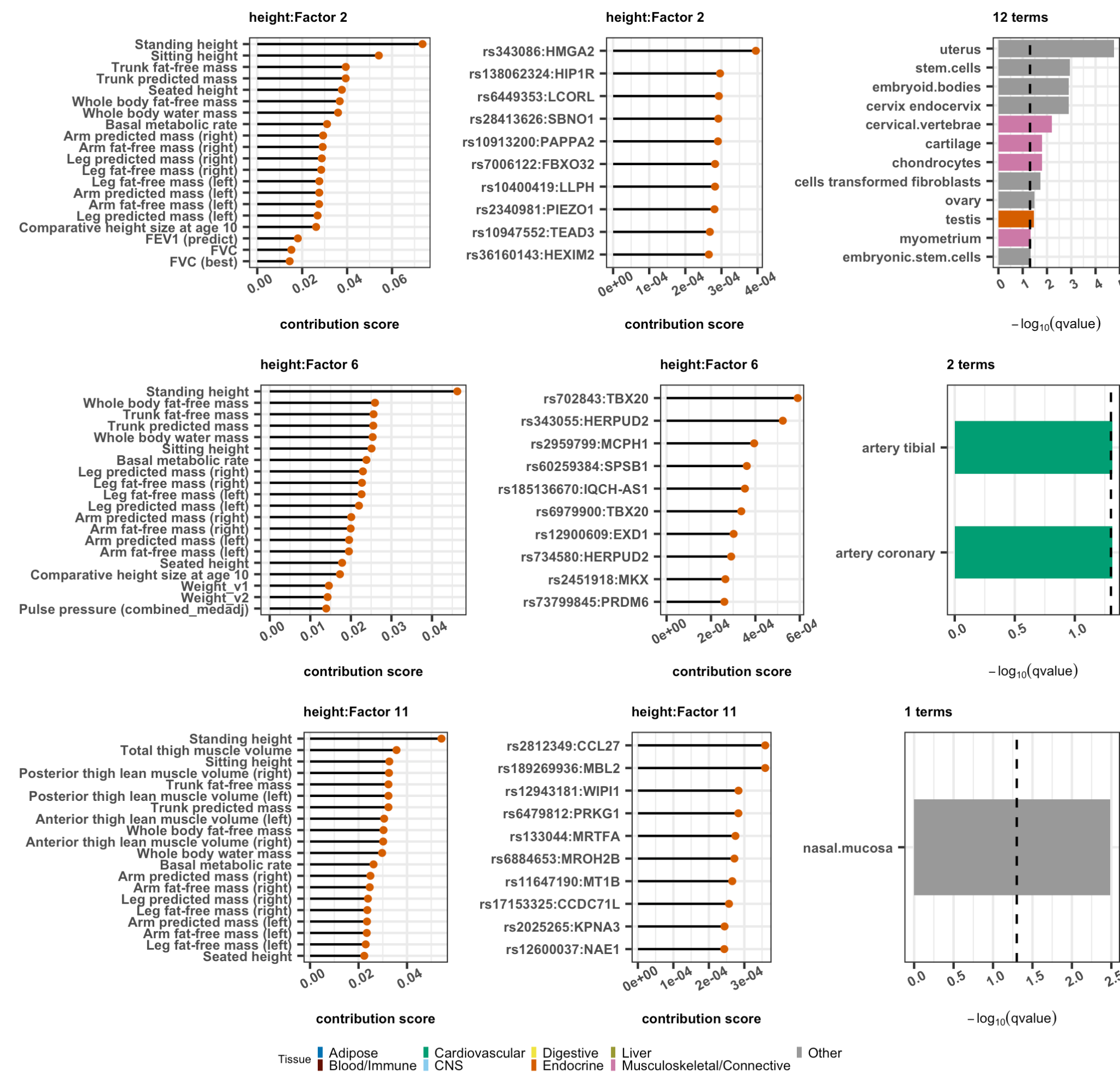

**Figure S14. Characterizing three leading factors in FactorGo for height.**

Results for factor 2, 6 and 11 (row) include 20 leading traits, 10 leading variants with closest gene, and enriched LDSC-SEG tissue or cell type. Total number enriched annotations are included in the title on the third column. Detailed results in **Table S4**.

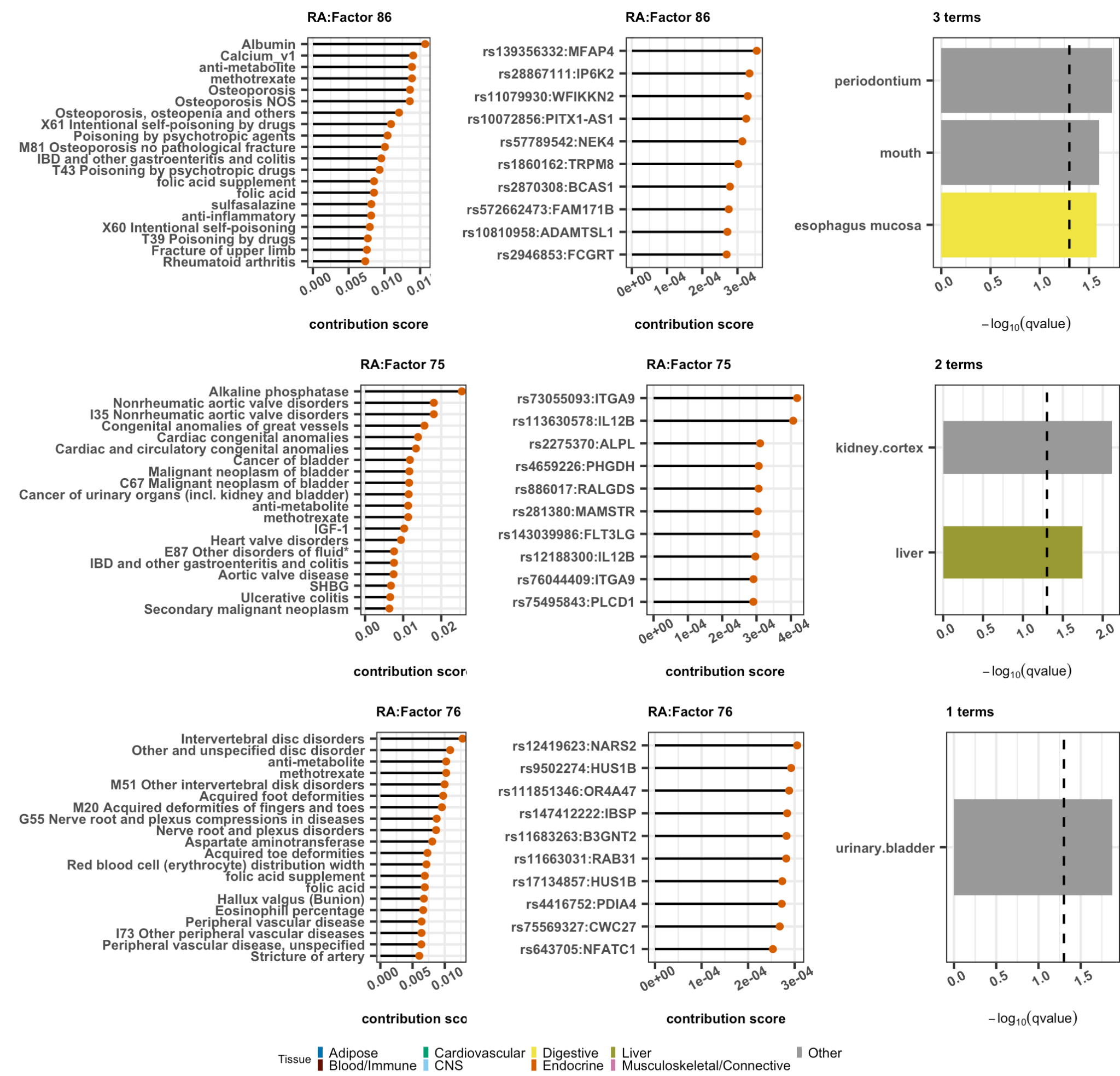

**Figure S15. Characterizing three leading factors in FactorGo for RA.**

Results for factor 86, 75 and 76 (row) include 20 leading traits, 10 leading variants with closest gene, and enriched LDSC-SEG tissue or cell type. Total number enriched annotations are included in the title on the third column. Detailed results in **Table S4**.

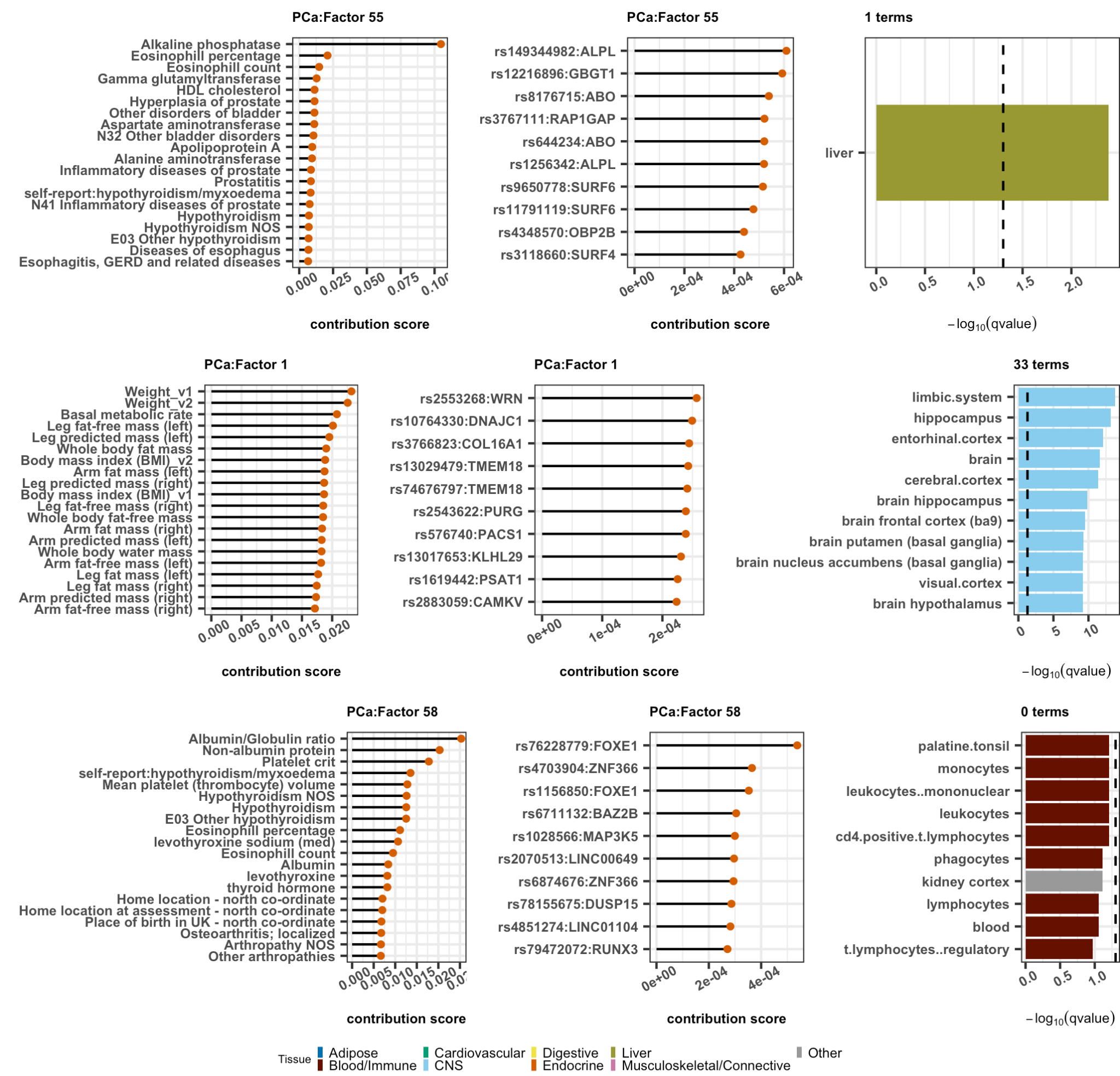

**Figure S16. Characterizing three leading factors in FactorGo for PCa.**

Results for factor 55, 1 and 58 (row) include 20 leading traits, 10 leading variants with closest gene, and enriched LDSC-SEG tissue or cell type. Total number enriched annotations are included in the title on the third column. Detailed result in **Table S4**.

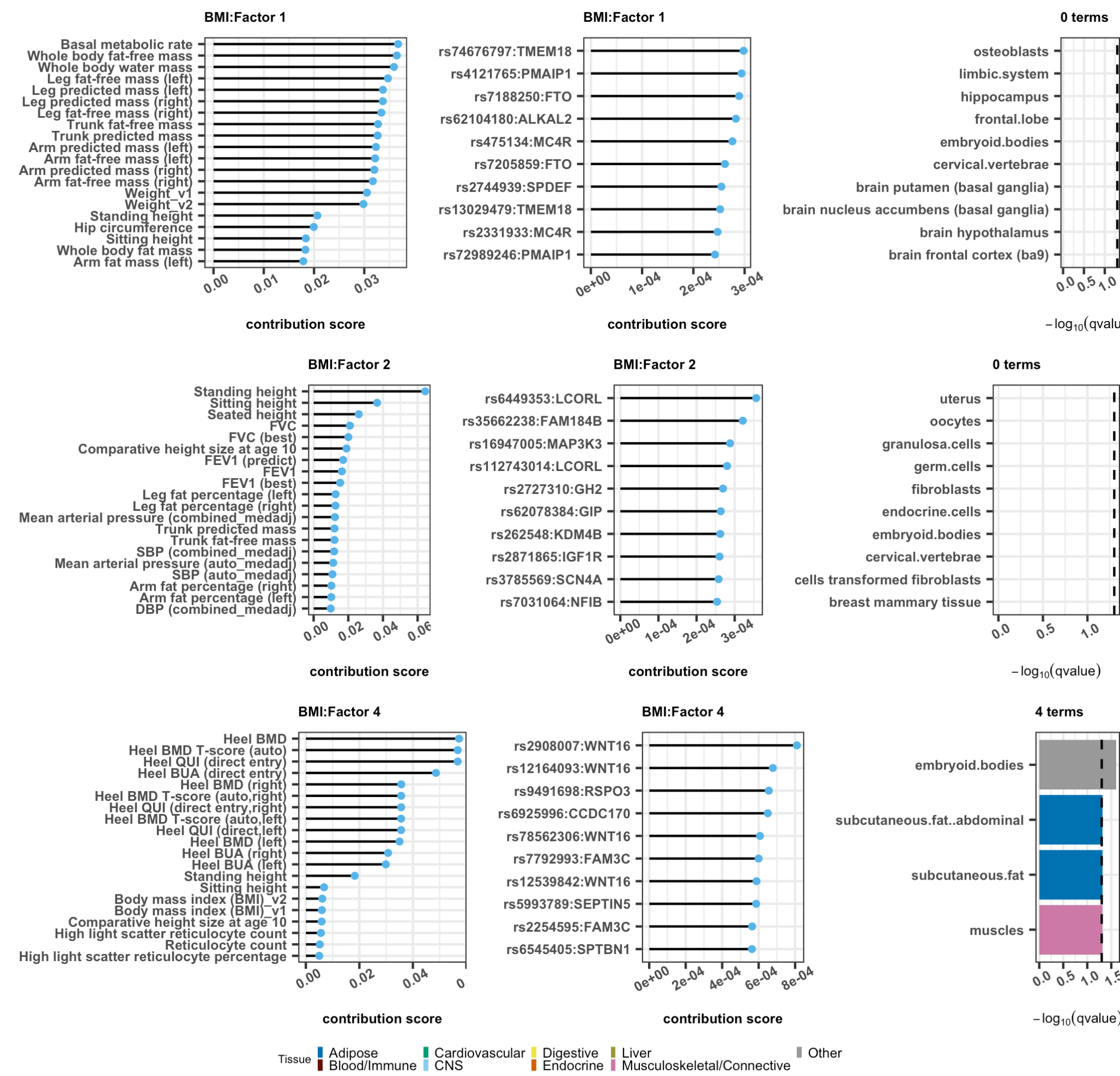

**Figure S17. Characterizing three leading factors in tSVD for BMI.**

Results for factor 1, 2 and 4 (row) include 20 leading traits, 10 leading variants with closest gene, and enriched LDSC-SEG tissue or cell type. Total number enriched annotations are included in the title on the third column. Detailed result in **Table S5**.

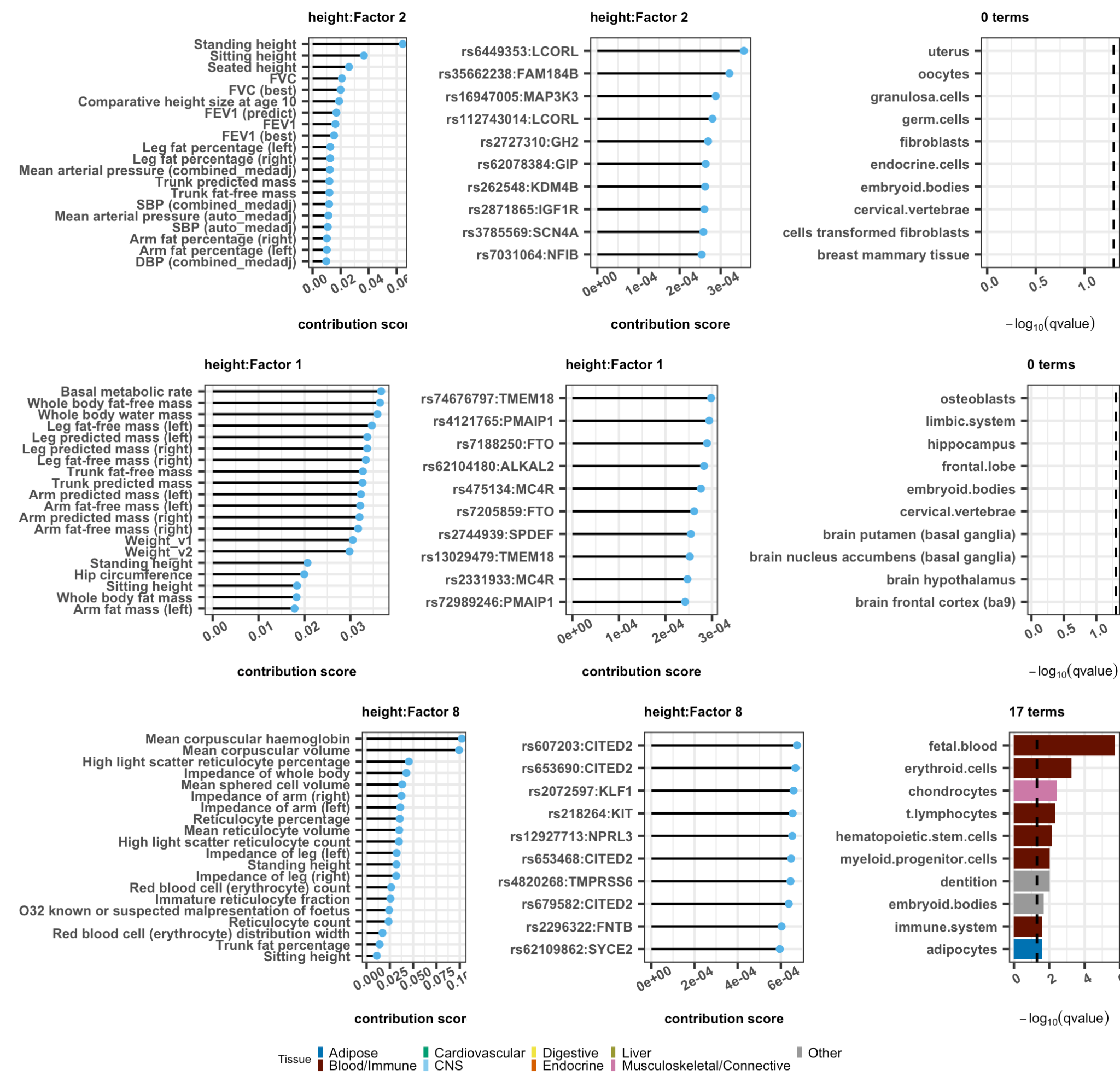

**Figure S18. Characterizing three leading factors in tSVD for height.**

Results for factor 2, 1 and 8 (row) include 20 leading traits, 10 leading variants with closest gene, and enriched LDSC-SEG tissue or cell type. Total number enriched annotations are included in the title on the third column. Detailed results in **Table S5**.

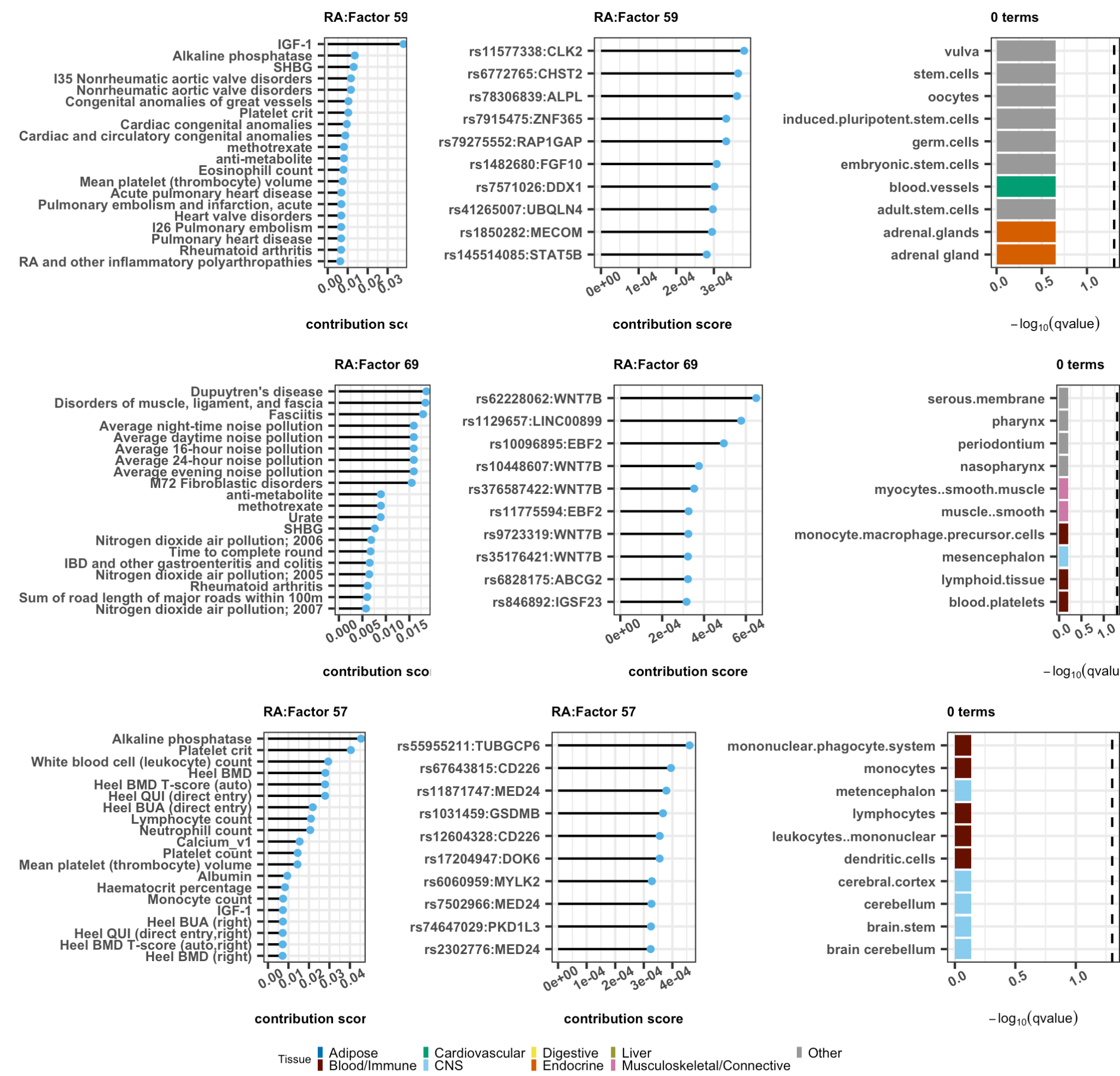

**Figure S19. Characterizing three leading factors in tSVD for RA.**

Results for factor 59, 69 and 57 (row) include 20 leading traits, 10 leading variants with closest gene, and enriched LDSC-SEG tissue or cell type. Total number enriched annotations are included in the title on the third column. Detailed results in **Table S5**.

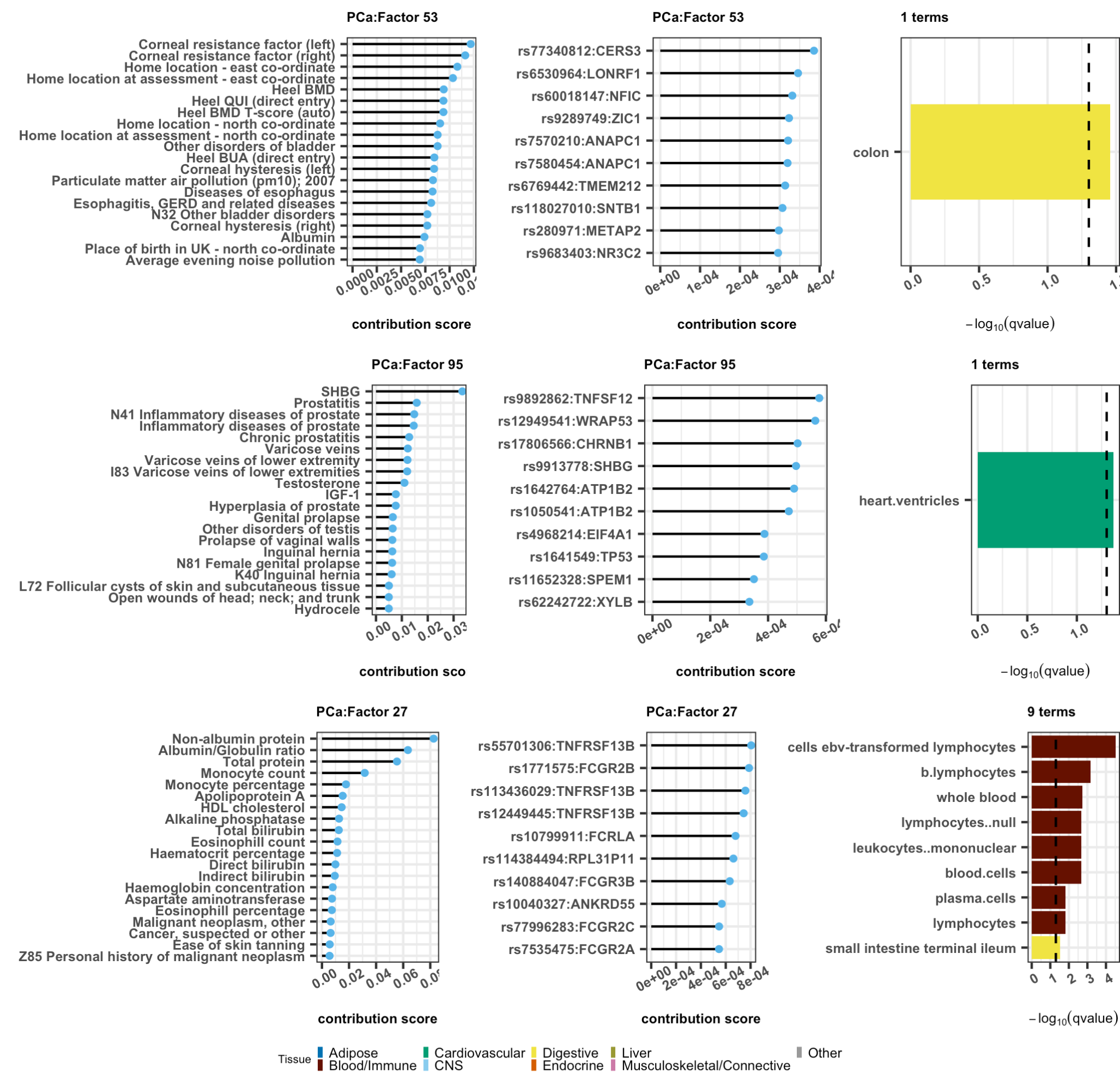

**Figure S20. Characterizing three leading factors in tSVD for PCa.**

Results for factor 53, 95 and 27 (row) include 20 leading traits, 10 leading variants with closest gene, and enriched LDSC-SEG tissue or cell type. Total number enriched annotations are included in the title on the third column. Detailed results in **Table S5**.

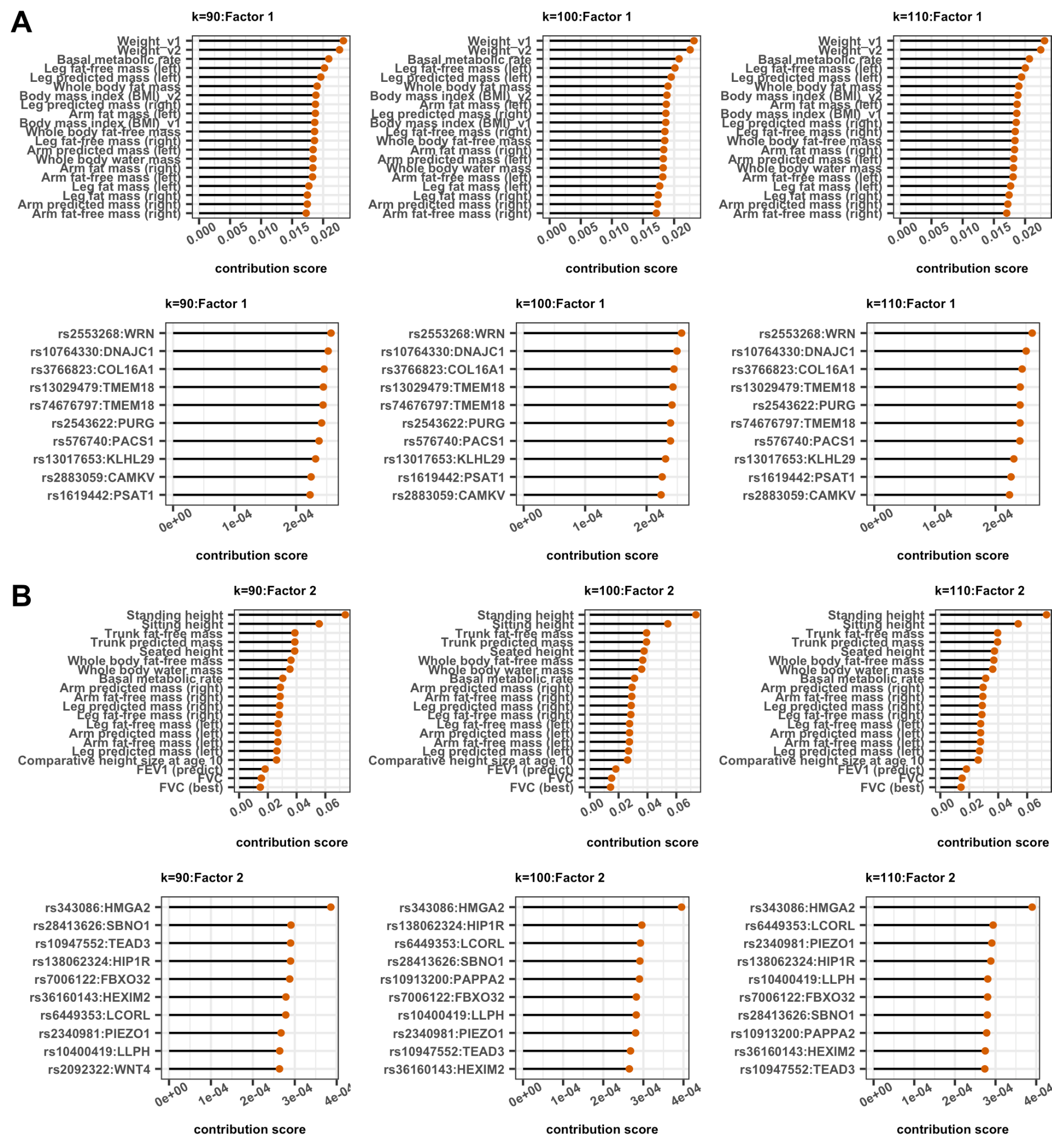

**Figure S21. Top two leading factors in FactorGo are robust to choices of k.**

**(A)** Compare 20 leading traits and 10 leading variants in factor 1 across $k=90,100,110$;

**(B)** Compare 20 leading traits and 10 leading variants in factor 2 across $k=90,100,110$.

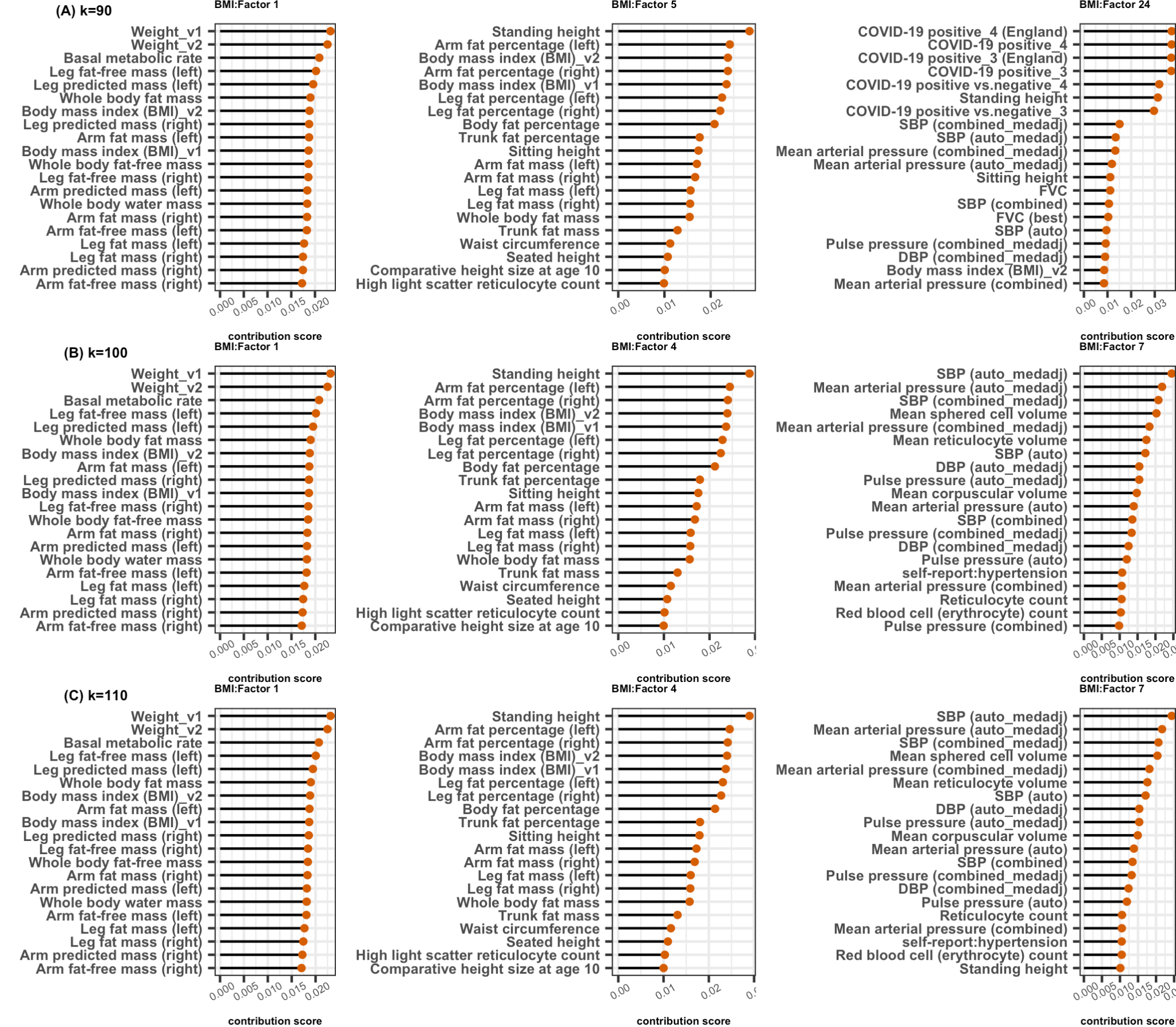

**Figure S22. Three leading factors for BMI in FactorGo are robust to choices of k.**

Compare 20 leading traits in three leading factors in FactorGo results using **(A)** $k=90$, **(B)** $k=100$, **(C)** $k=110$.

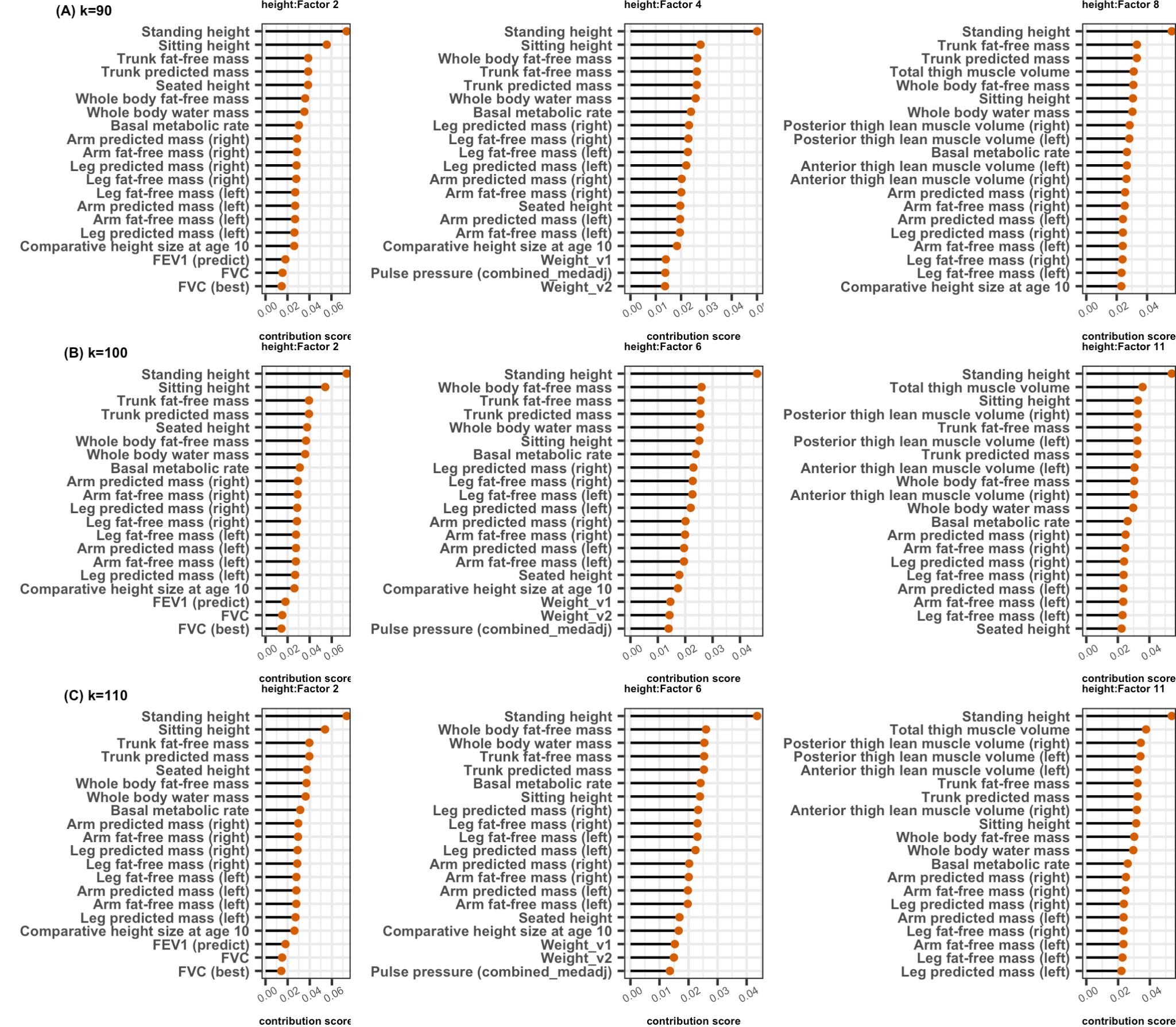

**Figure S23. Three leading factors for height in FactorGo are robust to choices of k.**

Compare 20 leading traits in three leading factors in FactorGo results using **(A)** $k=90$, **(B)** $k=100$, **(C)** $k=110$.

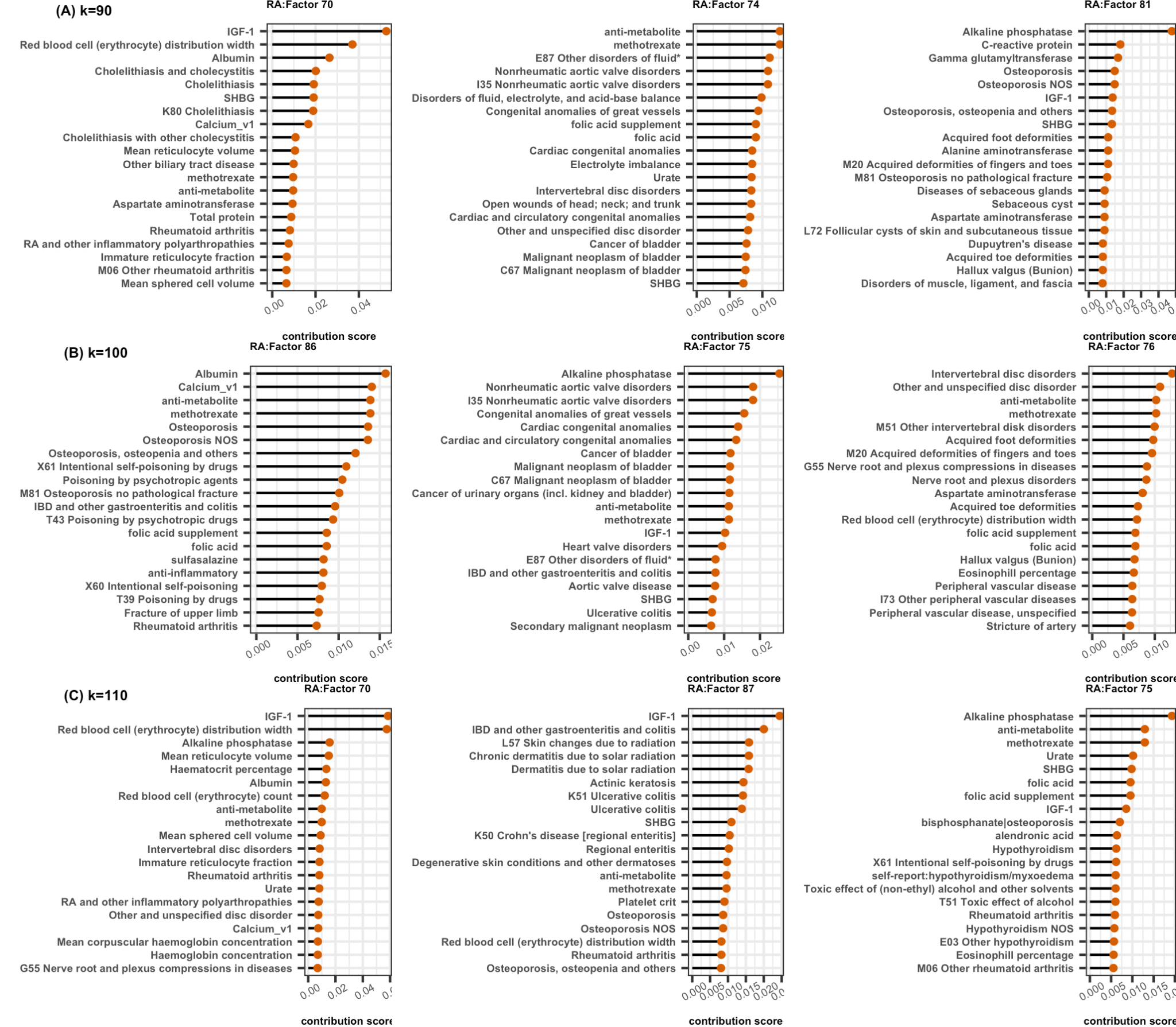

**Figure S24. Three leading factors for RA in FactorGo are overall robust to choices of k.**

Compare 20 leading traits in three leading factors in FactorGo results using **(A)** $k=90$, **(B)** $k=100$, **(C)** $k=110$.

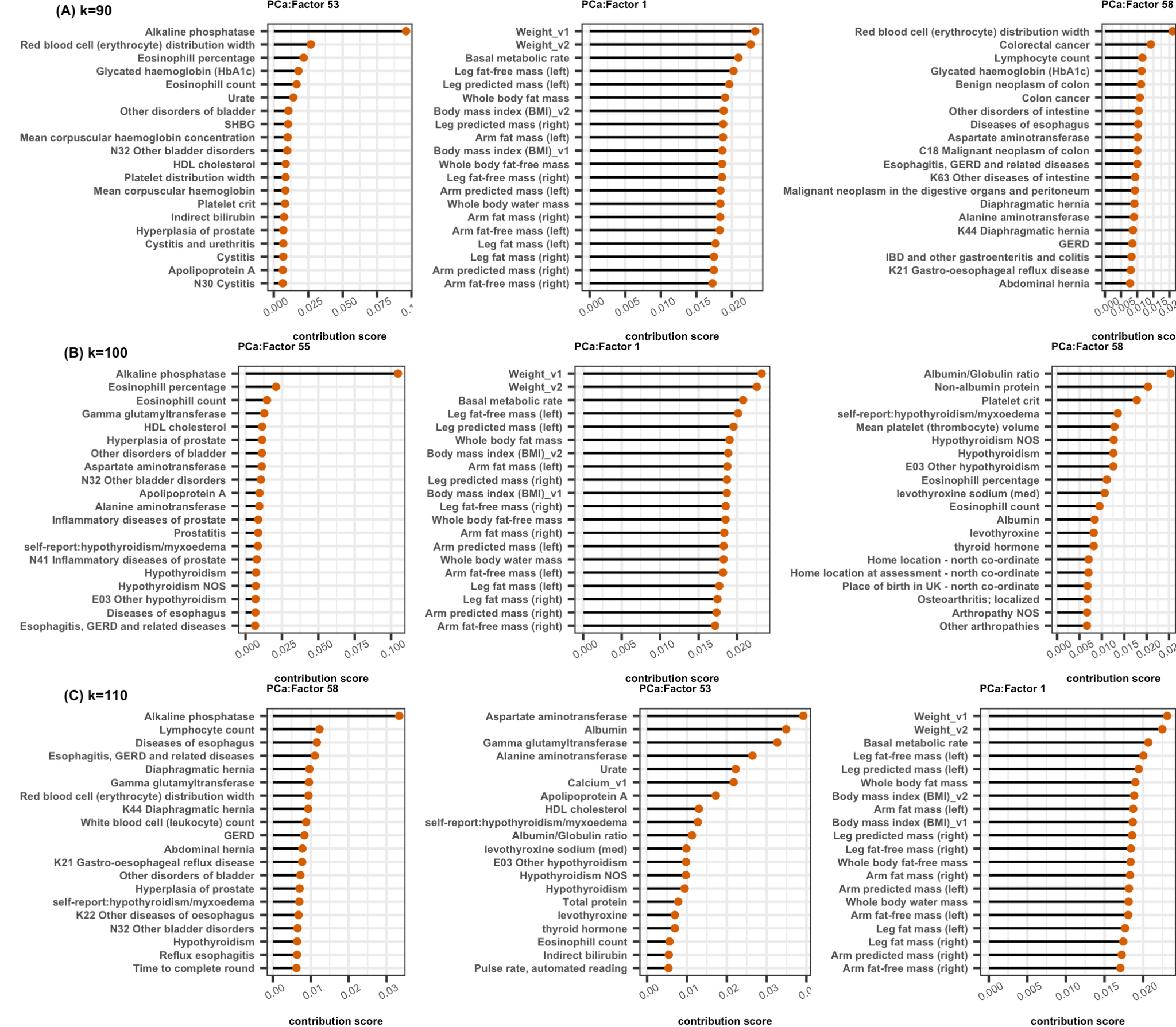

**Figure S25. Three leading factors for PCa in FactorGo are overall robust to choices of k.**

Compare 20 leading traits in three leading factors in FactorGo results using **(A)** $k=90$, **(B)** $k=100$, **(C)** $k=110$.

**Supplemental Tables**

**Table S1. Groups of 2,483 phenotypes**

| **Group** | **Type** | **Abbreviation** | **Number of traits** | **Example** | **UKB Field ID** |
| --- | --- | --- | --- | --- | --- |
| Disease | BIN | Disease (D) | 1130 | Type 2 diabetes | ICD10, phecode, self-reported 20002, COVID19 |
| Cancer | BIN | Cancer | 20 | Breast Cancer | self-reported 20001, phecode |
| Family history | BIN | Family history (FH) | 26 | Illness of mother, illness of father | 20107, 20110 |
| Treatment/Medication/Prescription | BIN | Medication/Prescription (MED) | 501 | Aspirin | 20003, 42039 harmonized |
| Physical measures | QT | Physical measures (PM) | 202 | Weight, Pulse rate | Category 100003, 100006,  derived variables |
| Mental health | QT | Mental Health (MENT) | 94 | General happiness, prospective memory | Category 100026, 100059, 136 |
| Biological samples (eg. assay) | QT | Assay | 67 | Cholesterol, Monocyte count | Category 100078 |
| Questionnaire  (eg.food intake, exercise, environment) | QT | Lifestyle and Exposure (LIFE) | 418 | Milk intake, Smoking, Physical activity | Category 100090, 100025, 113,123 |
| Misc | QT | Miscellaneous (MISC) | 25 | Birth weight, number of operations, home location |  |

**Table S2. 13 Body fat mass traits.**

Phenocode is a field ID described by UKB.

| **Phenocode** | **Description** |
| --- | --- |
| 22410 | Total trunk fat volume |
| 23099 | Body fat percentage |
| 23100 | Whole body fat mass |
| 23111 | Leg fat percentage (right) |
| 23112 | Leg fat mass (right) |
| 23115 | Leg fat percentage (left) |
| 23116 | Leg fat mass (left) |
| 23119 | Arm fat percentage (right) |
| 23120 | Arm fat mass (right) |
| 23123 | Arm fat percentage (left) |
| 23124 | Arm fat mass (left) |
| 23127 | Trunk fat percentage |
| 23128 | Trunk fat mass |

**Table S3. 5 Osteoporosis traits**

Phenocode is a field ID described by UKB.

| **Phenocode** | **Description** |
| --- | --- |
| M81 | M81 Osteoporosis without pathological fracture |
| 743 | Osteoporosis, osteopenia and pathological fracture |
| 743.1 | Osteoporosis |
| 743.11 | Osteoporosis NOS |
| 20002 | self-report:osteoporosis |
