## Supplemental Text 1 for "A scalable variational approach to characterize pleiotropic components across thousands of human diseases and complex traits using GWAS summary statistics"

### 1 Overview

Conventional factor analysis model decomposes the variance-covariance matrix of observed data into common variance due to shared latent factors and specific variance [1]. Both common and specific variance are estimated from data. To leverage the uncertainty estimates $SE^2$ in effect sizes from GWAS summary statistics, we extended the conventional factor analysis to use observed standard error as specific variance in Gaussian distribution. Taking a step further under reasonable assumptions, we simplified this SNP effect model to Z-score model (FactorGo). Under a Bayesian hierarchical model, FactorGo leverages variational inference to infer posterior moments of latent parameters. We applied a parameter expansion design to substantially accelerate convergence rate.

### 2 FactorGo model

#### 2.1 Notation

We denote matrices in uppercase bold (e.g., $\mathbf{X}$), vectors in lowercase bold (e.g., $\mathbf{x}$), and scalars in italicized lowercase (e.g., $x$). Further, we denote the $i^{th}$ column of matrix $\mathbf{X}$ as column-vector $\mathbf{x}_i$, and the $j^{th}$ row of matrix $\mathbf{X}$ as as column-vector $\mathbf{x}^j$. We denote the transpose of a matrix as $\mathbf{X}^\intercal$ and vector as $\mathbf{x}^\intercal$.

#### 2.2 Model

Let $\hat{\boldsymbol{\beta}}_i$ be a $p \times 1$ vector of effect size estimates at $p$ variants from GWAS trait $i$ and let $\hat{\boldsymbol{\Sigma}}_i$ represent the $p \times p$ diagonal matrix containing squared standard errors. We model the estimated SNP effects for trait $i$ as

$$\hat{\boldsymbol{\beta}}_i \sim \mathcal{N}(\mathbf{L}\mathbf{f}_i + \boldsymbol{\mu}, \boldsymbol{\tau}^{-1}\hat{\boldsymbol{\Sigma}}_i),$$

where $\mathbf{L}$ is a $p \times k$ factor loading matrix shared by all $n$ traits, $\mathbf{f}_i$ is the $k \times 1$ latent factor scores for trait $i$, $\boldsymbol{\mu}$ is a $p \times 1$ intercept vector. Conditional on shared loadings $\mathbf{L}$ and latent factors $\mathbf{f}_i$, we assume residuals are independent such that the off diagonals of $\hat{\boldsymbol{\Sigma}}_i$ are zeros. In practice, $\hat{\boldsymbol{\beta}}_i$ can be correlated due to linkage disequilibrium (i.e. LD) patterns, however we can perform LD-pruning to analyze a subset of variants such that LD is relatively minimal. Lastly, we let $\boldsymbol{\tau} = \sigma^{-2}$ capture any cross-study heterogeneity.

To simplify our model, we standardize effect sizes by pre-multiplying it with its standard errors such that

$$\hat{\mathbf{z}}_i \sim \mathcal{N}(\hat{\boldsymbol{\Sigma}}_i^{-1/2}(\mathbf{L}\mathbf{f}_i + \boldsymbol{\mu}), \boldsymbol{\tau}^{-1}\mathbf{I}_p).$$

where $\hat{\mathbf{z}}_i = \hat{\boldsymbol{\Sigma}}_i^{-1/2}\hat{\boldsymbol{\beta}}_i$. We note that standard errors for variant $j$ in study $i$ are proportional to $\sqrt{\frac{1}{N_i 2 f_{ij}(1-f_{ij})}}$, where $f_{ij}$ is the minor allele frequency at variant $j$ and $N_i$ is the $i^{th}$ GWAS total sample size. We assume that allele frequencies at variant $j$ are roughly constant across studies when the underlying population reflects similar ancestries. Thus this per-variant scaling term can be absorbed into the loading matrix $\mathbf{L}$ and intercept $\boldsymbol{\mu}$ giving,

$$\hat{\mathbf{z}}_i \sim \mathcal{N}(\sqrt{N_i}(\mathbf{L}\mathbf{f}_i + \boldsymbol{\mu}), \boldsymbol{\tau}^{-1}\mathbf{I}_p).$$

Similar to the Bayesian PCA approach proposed by Bishop 1999 [2], we impose full Bayesian treatment to this factor analysis model. The latent structure modeled by $\mathbf{L} = [\boldsymbol{\ell}^1, \dots, \boldsymbol{\ell}^p]^\intercal$, $\mathbf{F} = [\mathbf{f}_1, \dots, \mathbf{f}_n]$ and $\boldsymbol{\mu}$ has prior distributions as

$$\Pr(\boldsymbol{\mu}) = \mathcal{N}(\boldsymbol{\mu} \mid \mathbf{0}, \phi^{-1}\mathbf{I}_p)$$
$$\Pr(\mathbf{F}) = \prod_{i=1}^{n} \mathcal{N}(\mathbf{f}_i \mid \mathbf{0}, \mathbf{I}_k)$$
$$\Pr(\mathbf{L} \mid \boldsymbol{\alpha}) = \prod_{j=1}^{p} \mathcal{N}(\boldsymbol{\ell}^j \mid \mathbf{0}, \operatorname{diag}(\boldsymbol{\alpha}^{-1}))$$

To regularize model complexity, we put automatic relevance determination (ARD) priors on the loading matrix $\mathbf{L}$ such that less informative factors are shrunk towards zero. For each factor $q$, the ARD parameter $\boldsymbol{\alpha}_q$ is proportional to the inverse precision of that factor and modeled as Gamma distribution. The prior distributions are specified as follows:

$$\Pr(\boldsymbol{\alpha} \mid a_\alpha, b_\alpha) = \prod_{q=1}^{k} \Gamma(\boldsymbol{\alpha}_q \mid a_\alpha, b_\alpha)$$
$$\Pr(\boldsymbol{\tau} \mid a_\tau, b_\tau) = \Gamma(\boldsymbol{\tau} \mid a_\tau, b_\tau)$$

#### 2.3 Compare to tSVD

Truncated singular value decomposition (tSVD) is a reduced rank representation of original data matrix using result of SVD [3]. The full SVD decomposition for an observed Z-score summary statistics matrix is:

$$\hat{\mathbf{Z}}_{n\times p} = \mathbf{U}\mathbf{S}\mathbf{V}^T = \sum_{i=1}^{p} \mathbf{u}_i \mathbf{s}_i \mathbf{v}_i^T$$

Then tSVD has:

$$\hat{\mathbf{Z}}_{n\times p} \approx \sum_{i=1}^{k} \mathbf{u}_i \mathbf{s}_i \mathbf{v}_i^T$$

Unlike model-free tSVD, FactorGo appropriately accounts for the uncertainty in Z-scores due to differential power of GWAS studies and automatically infer model complexity. If $N_i$ is constant across studies and $\boldsymbol{\tau}^{-1}$ approaches 0, then we expect FactorGo produce similar result as tSVD.

#### 2.4 Variational Inference

##### 2.4.1 Overview

Considering the large number of parameters to estimate and its scalability to large dataset, we chose variational inference (VI) over other inference technique such as MCMC to infer posterior distribution of unknown parameters [4]. Unlike MCMC that aims to sample from true posterior distribution, VI converts this estimation problem to optimization problem. Given a choice of a tractable surrogate distribution $Q(\cdot)$ for the non-tractable true posterior, we solve for the optimal estimates to maximize the evidence lower bound (ELBO) of marginal log likelihood of data.

Let $\boldsymbol{\theta} = (\mathbf{L}, \mathbf{F}, \boldsymbol{\mu}, \boldsymbol{\alpha}, \boldsymbol{\tau})$ contains all unknown parameters, $\boldsymbol{\eta} = (a_\alpha, b_\alpha, a_\tau, b_\tau, \phi)$ be user specified hyperparameters and outcome Z-score data is $\hat{\mathbf{Z}}$. Suppose $Q(\boldsymbol{\theta})$ is any surrogate distribution for true posteriors, we can show the ELBO is a rigorous lower bound for marginal data likelihood by Janssen's inequality:

$$\begin{aligned}
\log P(\hat{\mathbf{Z}}) &= \log \int P(\hat{\mathbf{Z}}, \boldsymbol{\theta}) d\boldsymbol{\theta} \\
&= \log \int Q(\boldsymbol{\theta}) \frac{P(\hat{\mathbf{Z}}, \boldsymbol{\theta})}{Q(\boldsymbol{\theta})} d\boldsymbol{\theta} \\
&\geq \int Q(\boldsymbol{\theta}) \log \frac{P(\hat{\mathbf{Z}}, \boldsymbol{\theta})}{Q(\boldsymbol{\theta})} d\boldsymbol{\theta} \\
&= ELBO(Q)
\end{aligned}$$

The difference between $\log P(\hat{\mathbf{Z}})$ and lower bound $ELBO(Q)$ is called Kullback-Leibler (KL) divergence:

$$\begin{aligned}
\log P(\hat{\mathbf{Z}}) - ELBO(Q) &= E_Q(\log P(\hat{\mathbf{Z}})) - ELBO(Q) \\
&= \int Q(\boldsymbol{\theta}) \log P(\hat{\mathbf{Z}}) d\boldsymbol{\theta} - \int Q(\boldsymbol{\theta}) \log \frac{P(\hat{\mathbf{Z}}, \boldsymbol{\theta})}{Q(\boldsymbol{\theta})} d\boldsymbol{\theta} \\
&= \int Q(\boldsymbol{\theta})(\log P(\hat{\mathbf{Z}}) - \log \frac{P(\hat{\mathbf{Z}}, \boldsymbol{\theta})}{Q(\boldsymbol{\theta})}) d\boldsymbol{\theta} \\
&= \int Q(\boldsymbol{\theta}) \log \frac{Q(\boldsymbol{\theta})}{P(\boldsymbol{\theta} \mid \hat{\mathbf{Z}})} d\boldsymbol{\theta} \\
&= \mathrm{KL}[Q(\boldsymbol{\theta}) \parallel P(\boldsymbol{\theta} \mid \hat{\mathbf{Z}}, \boldsymbol{\eta})]
\end{aligned}$$

The relationship between these quantities is:

$$ELBO(Q) = \log P(\hat{\mathbf{Z}} \mid \boldsymbol{\eta}) - \mathrm{KL}[Q(\boldsymbol{\theta}) \parallel P(\boldsymbol{\theta} \mid \hat{\mathbf{Z}}, \boldsymbol{\eta})]$$

The *complete-data* likelihood $\mathcal{L}(\boldsymbol{\theta} \mid \hat{\mathbf{Z}}, \boldsymbol{\eta})$ is:

$$\mathcal{L}(\boldsymbol{\theta} \mid \hat{\mathbf{Z}}, \boldsymbol{\eta}) = \Pr(\boldsymbol{\tau} \mid a_\tau, b_\tau) \Pr(\boldsymbol{\alpha} \mid a_\alpha, b_\alpha) \Pr(\mathbf{L} \mid \boldsymbol{\alpha}) \Pr(\boldsymbol{\mu} \mid \phi) \prod_n \Pr(\hat{\mathbf{z}}_n \mid \boldsymbol{\mu}, \mathbf{f}_i, \mathbf{L}, \boldsymbol{\alpha}, \boldsymbol{\tau}, N_i) \Pr(\mathbf{f}_i)$$

Here we chose a fully factorizable distribution $Q$ from conjugate family such that:

$$Q(\mathbf{L}, \mathbf{F}, \boldsymbol{\alpha}, \boldsymbol{\tau}, \boldsymbol{\mu}) = Q(\mathbf{L}) Q(\mathbf{F}) Q(\boldsymbol{\alpha}) Q(\boldsymbol{\tau}) Q(\boldsymbol{\mu})$$

The solution for each parameter $\boldsymbol{\theta}_i$ is found by maximizing the the lower bound with respective to the following quantity:

$$Q(\boldsymbol{\theta}_i) \propto \mathbb{E}_{-i}[\log \mathcal{L}(\hat{\mathbf{Z}}, \boldsymbol{\theta})]$$

where the expectation is with respect to all parameters except $\boldsymbol{\theta}_i$.

Using completing squares, we can write out the posterior means and variances for multivariate Gaussian distribution or Gamma distribution. Here we provide solutions for each $Q$.

##### 2.4.2 $Q(\mathbf{F})$

$$
\begin{aligned}
Q(\mathbf{F}) &= \prod_{i=1}^{n} \mathcal{N}(\mathbf{f}_i \mid \mathbf{m}_{\mathbf{f}_i}, \mathbf{V}_{\mathbf{f}_i}) \text{ where} \\
\mathbf{V}_{\mathbf{f}_i} &= (\mathbf{I}_k + N_i \mathbb{E}[\boldsymbol{\tau}]\mathbb{E}[\mathbf{L}^\intercal \mathbf{L}])^{-1} \\
&= \left(\mathbf{I}_k + N_i \mathbb{E}[\boldsymbol{\tau}] \sum_{j=1}^{p} \mathbb{E}[\boldsymbol{\ell}^j \boldsymbol{\ell}^{j\intercal}]\right)^{-1} \\
&= \left(\mathbf{I}_K + N_i \mathbb{E}[\boldsymbol{\tau}] \sum_{j=1}^{p} (\mathbf{V}_{\boldsymbol{\ell}^j} + \mathbf{m}_{\boldsymbol{\ell}^j} \mathbf{m}_{\boldsymbol{\ell}^j}^\intercal)\right)^{-1} \\
\mathbf{m}_{\mathbf{f}_i} &= \sqrt{N_i} \mathbb{E}[\boldsymbol{\tau}] \mathbf{V}_{\mathbf{f}_i} \mathbb{E}[\mathbf{L}]^\intercal (\hat{\mathbf{z}}_i - \sqrt{N_i} \mathbb{E}[\boldsymbol{\mu}])
\end{aligned}
$$

### 2.4.3 $Q(\mu)$

$$
\begin{aligned}
Q(\boldsymbol{\mu}) &= \mathcal{N}(\boldsymbol{\mu} \mid \mathbf{m}_{\boldsymbol{\mu}}, \mathbf{V}_{\boldsymbol{\mu}}) \text{ where} \\
\mathbf{V}_{\boldsymbol{\mu}} &= (\phi + \mathbb{E}[\boldsymbol{\tau}] \sum_{i=1}^{n} N_i)^{-1} \mathbf{I}_p \\
\mathbf{m}_{\boldsymbol{\mu}} &= \mathbb{E}[\boldsymbol{\tau}] \mathbf{V}_{\boldsymbol{\mu}} \sum_{i=1}^{n} \sqrt{N_i} (\hat{\mathbf{z}}_i - \sqrt{N_i} \mathbb{E}[\mathbf{L}] \mathbb{E}[\mathbf{f}_i])
\end{aligned}
$$

##### 2.4.4 $Q(\mathbf{L})$

$$
\begin{aligned}
Q(\mathbf{L}) &= \prod_{j=1}^{p} \mathcal{N}(\boldsymbol{\ell}^j \mid \mathbf{m}_{\boldsymbol{\ell}^j}, \mathbf{V}_{\boldsymbol{\ell}^j}) \text{ where} \\
\mathbf{V}_{\boldsymbol{\ell}^j} &= \left(\operatorname{diag}(\mathbb{E}[\boldsymbol{\alpha}]) + \mathbb{E}[\boldsymbol{\tau}] \sum_{i=1}^{n} N_i \mathbb{E}[\mathbf{f}_i \mathbf{f}_i^\intercal]\right)^{-1} \\
&= \left(\operatorname{diag}(\mathbb{E}[\boldsymbol{\alpha}]) + \mathbb{E}[\boldsymbol{\tau}] \sum_{i=1}^{n} N_i (\mathbf{V}_{\mathbf{f}_i} + \mathbf{m}_{\mathbf{f}_i} \mathbf{m}_{\mathbf{f}_i}^\intercal)\right)^{-1} \\
\mathbf{m}_{\boldsymbol{\ell}^j} &= \mathbb{E}[\boldsymbol{\tau}] \mathbf{V}_{\boldsymbol{\ell}^j} \left(\sum_{i=1}^{n} \sqrt{N_i} \mathbb{E}[\mathbf{f}_i] (\hat{\mathbf{z}}_{ij} - \sqrt{N_i} \mathbb{E}[\boldsymbol{\mu}_j])\right)
\end{aligned}
$$

###### 2.4.5 $Q(\alpha)$

$$
\begin{aligned}
Q(\boldsymbol{\alpha}) &= \prod_k \Gamma(\boldsymbol{\alpha}_k \mid \tilde{a}_\alpha, \tilde{b}_{\alpha k}) \text{ where} \\
\tilde{a}_\alpha &= a_\alpha + \frac{p}{2} \\
\tilde{b}_{\alpha k} &= b_\alpha + \frac{\mathbb{E}[\boldsymbol{\ell}_k^\intercal \boldsymbol{\ell}_k]}{2} = b_\alpha + \frac{\operatorname{diag}(\mathbb{E}[\mathbf{L}^\intercal \mathbf{L}])}{2} \\
&= b_\alpha + \frac{1}{2} \sum_{j=1}^{p} \mathbb{E}[\boldsymbol{\ell}^j \boldsymbol{\ell}^{j\intercal}]_{(kk)} \\
&= b_\alpha + \frac{1}{2} \sum_{j=1}^{p} [\mathbf{V}_{\boldsymbol{\ell}^j} + \mathbf{m}_{\boldsymbol{\ell}^j} \mathbf{m}_{\boldsymbol{\ell}^j}^\intercal]_{(kk)}
\end{aligned}
$$

###### 2.4.6 $Q(\tau)$

$$
\begin{aligned}
Q(\boldsymbol{\tau}) &= \Gamma(\boldsymbol{\tau} \mid \tilde{a}_\tau, \tilde{b}_\tau) \text{ where} \\
\tilde{a}_\tau &= a_\tau + \frac{np}{2} \\
\tilde{b}_\tau &= b_\tau + \frac{1}{2} \sum_{i=1}^{n} \mathbb{E}\|\hat{\mathbf{z}}_i - \sqrt{N_i}\mathbf{L}\mathbf{f}_i - \sqrt{N_i}\boldsymbol{\mu}\|^2
\end{aligned}
$$

###### 2.4.7 ELBO

After each iteration, calculate ELBO by $\mathbb{E}[\log \Pr(\hat{\mathbf{Z}} \mid \boldsymbol{\theta}, \boldsymbol{\eta})] - \mathrm{KL}[Q(\boldsymbol{\theta}) \parallel \Pr(\boldsymbol{\theta} \mid \boldsymbol{\eta})]$, where KL term can be calculate separately for each parameter.

Data likelihood term:

$$
\mathbb{E}[\log \Pr(\hat{\mathbf{Z}} \mid \boldsymbol{\theta}, \boldsymbol{\eta})] = \mathbb{E}\left[\log \prod_{i=1}^{n} \mathcal{N}(\sqrt{N_i}\mathbf{L}\mathbf{f}_i + \sqrt{N_i}\boldsymbol{\mu}, \boldsymbol{\tau}^{-1}) \mid \boldsymbol{\theta}, \boldsymbol{\eta}\right]
$$

**L**:

$$
\begin{aligned}
\mathbb{E}[\log Q(\boldsymbol{\ell}^j)] &= \mathbb{E}\left[-\frac{k}{2}\log 2\pi - \frac{1}{2}\log|\mathbf{V}_{\boldsymbol{\ell}^j}| - \frac{1}{2}(\boldsymbol{\ell}^j - \mathbf{m}_{\boldsymbol{\ell}^j})^\intercal \mathbf{V}_{\boldsymbol{\ell}^j}^{-1}(\boldsymbol{\ell}^j - \mathbf{m}_{\boldsymbol{\ell}^j}) \mid \boldsymbol{\theta}\right] \\
\mathbb{E}[\log \Pr(\boldsymbol{\ell}^j \mid \boldsymbol{\eta})] &= \mathbb{E}\left[-\frac{k}{2}\log 2\pi - \frac{1}{2}\log|\operatorname{diag}(\boldsymbol{\alpha}^{-1})| - \frac{1}{2}(\boldsymbol{\ell}^{j\intercal}\operatorname{diag}(\boldsymbol{\alpha})\boldsymbol{\ell}^j) \mid \boldsymbol{\eta}\right] \\
\mathrm{KL}[Q(\mathbf{L}) \parallel \Pr(\mathbf{L} \mid \boldsymbol{\eta})] &= \sum_{j=1}^{p} \mathbb{E}[\log Q(\boldsymbol{\ell}^j)] - \mathbb{E}[\log \Pr(\boldsymbol{\ell}^j \mid \boldsymbol{\eta})]
\end{aligned}
$$

**F**:

$$
\mathrm{KL}[Q(\mathbf{F}) \parallel \Pr(\mathbf{F} \mid \boldsymbol{\eta})] = \sum_{i=1}^{n} \mathbb{E}[\log Q(\mathbf{f}_i)] - \mathbb{E}[\log \Pr(\mathbf{f}_i \mid \boldsymbol{\eta})]
$$

$\boldsymbol{\mu}$:

$$\mathrm{KL}[Q(\boldsymbol{\mu}) \parallel \Pr(\boldsymbol{\mu} \mid \boldsymbol{\eta})] = \mathbb{E}[\log Q(\boldsymbol{\mu})] - \mathbb{E}[\log \Pr(\boldsymbol{\mu} \mid \boldsymbol{\eta})]$$

$\boldsymbol{\alpha}$:

$$\begin{aligned}
\mathbb{E}[\log \Gamma(\boldsymbol{\alpha}_k \mid \tilde{a}_\alpha, \tilde{b}_\alpha)] &= \tilde{a}_\alpha \log(\tilde{b}_\alpha) + (\tilde{a}_\alpha - 1)\mathbb{E}[\log \boldsymbol{\alpha}_k \mid \tilde{a}_\alpha, \tilde{b}_\alpha] - \tilde{b}_\alpha \mathbb{E}[\boldsymbol{\alpha}_k \mid \tilde{a}_\alpha, \tilde{b}_\alpha] - \log \Gamma(\tilde{a}_\alpha) \\
\mathbb{E}[\log \Gamma(\boldsymbol{\alpha}_k \mid a_\alpha, b_\alpha)] &= a_\alpha \log(b_\alpha) + (a_\alpha - 1)\mathbb{E}[\log \boldsymbol{\alpha}_k \mid \tilde{a}_\alpha, \tilde{b}_\alpha] - b_\alpha \mathbb{E}[\boldsymbol{\alpha}_k \mid \tilde{a}_\alpha, \tilde{b}_\alpha] - \log \Gamma(a_\alpha) \\
\mathrm{KL}[Q(\boldsymbol{\alpha}) \parallel \Pr(\boldsymbol{\alpha} \mid \boldsymbol{\eta})] &= \sum_k \mathbb{E}[\log \Gamma(\boldsymbol{\alpha}_k \mid \tilde{a}_\alpha, \tilde{b}_\alpha)] - \mathbb{E}[\log \Gamma(\boldsymbol{\alpha}_k \mid a_\alpha, b_\alpha)]
\end{aligned}$$

$\boldsymbol{\tau}$:

$$\begin{aligned}
\mathbb{E}[\log \Gamma(\boldsymbol{\tau} \mid \tilde{a}_\tau, \tilde{b}_\tau)] &= \tilde{a}_\tau \log(\tilde{b}_\tau) + (\tilde{a}_\tau - 1)\mathbb{E}[\log \boldsymbol{\tau} \mid \tilde{a}_\tau, \tilde{b}_\tau] - \tilde{b}_\tau \mathbb{E}[\boldsymbol{\tau} \mid \tilde{a}_\tau, \tilde{b}_\tau] - \log \Gamma(\tilde{a}_\tau) \\
\mathbb{E}[\log \Gamma(\boldsymbol{\tau} \mid a_\tau, b_\tau)] &= a_\tau \log(b_\tau) + (a_\tau - 1)\mathbb{E}[\log \boldsymbol{\tau} \mid \tilde{a}_\tau, \tilde{b}_\tau] - b_\tau \mathbb{E}[\boldsymbol{\tau} \mid \tilde{a}_\tau, \tilde{b}_\tau] - \log \Gamma(a_\tau) \\
\mathrm{KL}[Q(\boldsymbol{\tau}) \parallel \Pr(\tau \mid \boldsymbol{\eta})] &= \mathbb{E}[\log \Gamma(\boldsymbol{\tau} \mid \tilde{a}_\tau, \tilde{b}_\tau)] - \mathbb{E}[\log \Gamma(\boldsymbol{\tau} \mid a_\tau, b_\tau)]
\end{aligned}$$

#### 2.5 Parameter expansion

The convergence of FactorGo under the above model can be slow because $\mathbf{L}$ and $\mathbf{F}$ are strongly coupled in the model, whereas vectors in $\mathbf{L}$ and $\mathbf{F}$ are assumed to be independent a posteriori for computational convenience. To speed up inference, we applied a parameter expansion method proposed for variational Bayesian factor analysis specifically [5]. The general idea is to introduce auxilliary parameter for bias $\mathbf{b}$ and $\mathbf{R}$ in the posterior distribution that are optimized during inference. After each iteration step, we can jointly update the parameters in $\mathbf{L}, \mathbf{F}, \boldsymbol{\alpha}$. Here we provided the updating rule under this transformation method:

1. First remove bias between $\mathbf{F}$ and $\boldsymbol{\mu}$

$$\begin{aligned}
Q^*(\mathbf{F}) &= \prod_{i=1}^{n} \mathcal{N}(\mathbf{f}_i \mid \mathbf{m}_{\mathbf{f}_i} - \mathbf{b}, \mathbf{V}_{\mathbf{f}_i}) \\
Q^*(\boldsymbol{\mu}) &= \mathcal{N}(\boldsymbol{\mu} \mid \mathbf{m}_{\boldsymbol{\mu}} + \mathbf{L}\mathbf{b}, \mathbf{V}_{\boldsymbol{\mu}})
\end{aligned}$$

where

$$\begin{aligned}
\mathbf{b} &= (\sum_{i=1}^{n} \Psi_i)^{-1} (\sum_{i=1}^{n} \Psi_i \mathbf{m}_{\mathbf{f}_i}) \\
\Psi_i &= N_i \mathbb{E}[\boldsymbol{\tau}] p \mathbf{V}_{\boldsymbol{\ell}^j} + \mathbf{I}_k
\end{aligned}$$

2. Then rotate latent subspace

$$\begin{aligned}
Q^*(\mathbf{L}) &= \prod_{j=1}^{p} \mathcal{N}(\boldsymbol{\ell}^j \mid \mathbf{R}^\intercal \mathbf{m}_{\boldsymbol{\ell}^j}, \mathbf{R}^\intercal \mathbf{V}_{\boldsymbol{\ell}^j} \mathbf{R}) \\
Q^*(\mathbf{F}) &= \prod_{i=1}^{n} \mathcal{N}(\mathbf{f}_i \mid \mathbf{R}^{-1} \mathbf{m}_{\mathbf{f}_i}, \mathbf{R}^{-1} \mathbf{V}_{\mathbf{f}_i} \mathbf{R}^{-\intercal}) \\
Q^*(\boldsymbol{\alpha}) &= \prod_k \Gamma(\boldsymbol{\alpha}_k \mid \tilde{a}_\alpha, b_\alpha + \frac{1}{2} \mathrm{diag}(\mathbf{R}^\intercal \mathbb{E}_Q[\mathbf{L}^\intercal \mathbf{L}] \mathbf{R}))
\end{aligned}$$

The optimal rotation matrix $\mathbf{R}$ can be found by following steps:
let $\mathbf{R} = \mathbf{U}\boldsymbol{\Lambda}\mathbf{V}$, then $\mathbf{U}$ and $\boldsymbol{\Lambda}$ are found by eigen-decomposition of:

$$\frac{1}{n}\mathbb{E}_Q[\mathbf{F}\mathbf{F}^\intercal] = \mathbf{U}\boldsymbol{\Lambda}^2\mathbf{U}^\intercal$$

$\mathbf{V}$ is found by eigen-decomposition of:

$$\boldsymbol{\Lambda}\mathbf{U}^\intercal\mathbb{E}_Q[\mathbf{L}^\intercal\mathbf{L}]\mathbf{U}\boldsymbol{\Lambda} = \mathbf{V}\mathbf{D}\mathbf{V}^\intercal$$

##### 2.5.1 FactorGo algorithm

**Algorithm 1:** FactorGo with parameter expansion design

Input: GWAS Z-score summary data and sample size
Initialize: $a_\alpha = b_\alpha = a_\tau = b_\tau = \phi = 10^{-5}, \mathbb{E}[\boldsymbol{\ell}^j] = \mathbf{0}, \mathbb{V}(\boldsymbol{\ell}^j) = \mathbf{I}$ for all $j \in [p], ELBO_0 = 0$
**while** $ELBO_i - ELBO_{i-1} > 0.001$ *or* $i \leq itr_{max}$ **do**
    update $\mathbf{m}_\mathbf{F}, \mathbf{V}_\mathbf{F}$
    update $\mathbf{m}_{\boldsymbol{\mu}}, \mathbf{V}_{\boldsymbol{\mu}}$
    update $\mathbf{m}_\mathbf{L}, \mathbf{V}_\mathbf{L}$
    update $\mathbf{m}_{\boldsymbol{\alpha}}, \mathbf{V}_{\boldsymbol{\alpha}}$
    find optimal $\mathbf{b}, \mathbf{R}$ for transformation and update above parameters
    update $\mathbf{m}_{\boldsymbol{\tau}}, \mathbf{V}_{\boldsymbol{\tau}}$
    Calculate $ELBO$
**end**
Output: return posterior mean and variance of $\boldsymbol{\theta} = (\mathbf{L}, \mathbf{F}, \boldsymbol{\mu}, \boldsymbol{\alpha}, \boldsymbol{\tau})$.

#### 2.6 Calculate variance explained $R^2$ in observed data

Here we provide formula to calculate variance explained by each factor $R_k^2$. For $k^{th}$ factor, and $i^{th}$ study, let $\hat{\mathbf{z}}_i'$ be fitted Z-score value by all factors and $\hat{\mathbf{z}}_i^{(k)'}$ be fitted value for $k^{th}$ factor only:

$$\begin{aligned}
\hat{\mathbf{z}}_i' &= \sqrt{N_i}\mathbb{E}[\mathbf{L}]\mathbb{E}[\mathbf{f}_i] = \sqrt{N_i}\sum_k \mathbb{E}[\mathbf{f}_{ik}]\mathbb{E}[\boldsymbol{\ell}_k] \\
\hat{\mathbf{z}}_i^{(k)'} &= \sqrt{N_i}\mathbb{E}[\mathbf{f}_{ik}]\mathbb{E}[\boldsymbol{\ell}_k]
\end{aligned}$$

Then calculate $R_k^2$ using total residual error and total variance, where $\sigma^2$ is canceled:

$$\begin{aligned}
SSE_k &= \sum_i (\hat{\mathbf{z}}_i - \hat{\mathbf{z}}_i^{(k)'})^\intercal \sigma^{-2} (\hat{\mathbf{z}}_i - \hat{\mathbf{z}}_i^{(k)'}) \\
TSS &= \sum_i \hat{\mathbf{z}}_i^\intercal \sigma^{-2} \hat{\mathbf{z}}_i \\
R_k^2 &= 1 - \frac{SSE_k}{TSS}
\end{aligned}$$
