## Supplemental Text 2 for "A scalable variational approach to characterize pleiotropic components across thousands of human diseases and complex traits using GWAS summary statistics"

**Characterizing second and third leading factors for focal traits in FactorGo**

*BMI*

The next two leading factors for BMI identified its shared biology with pharynx and digestive system respectively (squared cosine score: 17.66%, 2.86%). Factor 4 characterized standing height (2.88%) and body fat measures (cumulative 22.73% across 13 traits; **Figure S13, Table S2**). Two leading variants were proximal to *PDE10A* (rs9459529:T>C: 0.030%) and *IGF1* (rs1113483:T>C: 0.029%), both of which play roles in energy homeostasis and obesity etiology^1,2^ . Factor 4 was enriched with genes specifically expressed in pharynx, which could reflect the negative association between upper airway size and body fat distribution^3^. Factor 7 characterized blood pressure traits that are strongly associated with BMI (25.12% across 25 traits; **Figure S13**)^4^. It was enriched with subcutaneous fat and digestive systems such as the colon and intestines, which supports obesity as a risk factor for colorectal cancer^5^.

*Height*

The next two leading factors for standing height identified its shared biology with cardiovascular and immunity respectively (squared cosine score: 9.64%, 8.51%). Driven by variant proximal to *TBX20* associated with heart growth (rs702843:G>C: 0.059%)^6^, factor 6 exhibited enrichment in coronary arteries (**Figure S14**), which is consistent with previous findings of enrichment in coronary tissues in height associated variants^7,8^. Factor 11 was driven by variants closest to *CCL27* and *MBL2* (rs2812349:T>C: 0.036%; rs189269936:C>G: 0.036%), both of which are involved in the immune system^9,10^. This was supported by its enrichment in nasal mucosa harboring diverse immune cells. Although the relationship between the immune system and human growth is unclear, it has been shown there is an energetic tradeoff between immune function and growth in the Amazonian^11^.

*Rheumatoid arthritis*

The next two leading factors for RA identified its shared biology with kidney, liver and urinary bladder (squared cosine score: 6.01%, 5.75%). Factor 75 characterized alkaline phosphatase (2.54%), cardiac (1.81%) and bladder disease (1.16%; **Figure S15**). The leading variants were close to *ITGA9* (rs73055093:C>T: 0.042%) and *IL12B* (rs113630578: A>G: 0.041%), both of which are important for the immune system. This factor showed enrichment in the kidney cortex and liver, both of which contain high levels of ALP enzymes. This can reflect the comorbidity of RA involved with liver and kidney disease^12,13^. Factor 76 characterized traits involved in the central nervous system such as intervertebral disc disorders (1.28%) and peripheral disorders such as foot deformities (0.97%; **Figure S15**). Its enrichment in the urinary bladder could reflect the shared symptoms of general reactive arthritis.

*Prostate cancer*

The next two leading factors for PCa identified BMI and hormonal disorder associated with prostate cancer respectively (squared cosine score: 7.71%, 6.37%). Since factor 2 was identified as BMI factor (see **Result**), this supported the impact of obesity on prostate cancer progression due to inflammation and metabolic mechanisms^14,15^. Driven by *FOXE1* associated with thyroid morphogenesis (**Figure S16**)^16^, factor 58 characterized blood-related traits such as albumin/globulin ratio (2.53%), platelet crit (1.78%) and other hormonal disorders such as hypothyroidism (1.26%), suggesting the shared mechanisms of PCa involved with hormones.
